# Human mobility data from the BBC Pandemic project

**DOI:** 10.1101/2021.02.19.21252079

**Authors:** Andrew JK Conlan, Petra Klepac, Adam J Kucharski, Stephen Kissler, Maria L Tang, Hannah Fry, Julia R Gog

## Abstract

We present human mobility data for the United Kingdom collected from the “BBC Pandemic”, a public science project linked to the BBC Four television documentary of the same name. Mobile phone GPS trajectories submitted by users and collected over a 24 hour period were aggregated to construct anonymised origin-destination flux matrices at the local administrative district (LAD). We use these data to explore how mobility patterns change with age and employment status - unique stratifications that are not available from other publicly and privately held mobility data sets. We validate the consistency of the aggregated BBC mobility data set against census workflow data and illustrate how the systematic differences in mobility rates with age affect the risk and pattern of transmission between regions with 18-30 year old’s contributing the greatest risk of transmission to adjacent regions, but older 60-100 years playing the most important role in more remote low-density locations.

## 1 Introduction

The spatial dynamics of human mobility are a key mechanism driving the local persistence of endemic infections [9, 10, 24, 18] and limiting the rate of invasion of novel pathogens [31]. Over the past twenty years there has been an rapid expansion in the type of data and methods available to directly track or infer patterns of human mobility [8] from traditional sources such as census data, tracking bank notes [14], the frequency of posting on social media [36], mobile phone records [23] through to direct tracking using the global positioning system (GPS) [48]. Mobile phone GPS data are perhaps the most accurate and powerful tool to measure human mobility - as demonstrated most recently by the real time analysis of the effectiveness of lockdown procedures during the Covid-19 pandemic [32, 46, 49]. However, mobile phone data sets are privately held by mobile network operators and – with a few notable exceptions [48, 37, 26] – rarely shared publicly which is a barrier to reproducibility and open science [35]. Hence, spatial epidemic models still often rely on traditional sources of mobility data such as census workflows for commuters [7, 28, 15, 5]. Given the importance of protecting individuals identities, mobile phone data shared privately to researchers by network operators also lacks key meta-data on users such as gender, age and employment status which are likely to affect patterns of mobility [13].

To address these limitations, the BBC Pandemic project recruited over 86,000 participants in the United Kingdom between September 2017 and December 2018 as part of a public science project linked to a BBC Four documentary [31]. Here we present and analyse mobility data from 43,291 participants aggregated to the level of the observed flux between local administrative districts and stratified by age and employment status. The only other open source mobility data currently available for the United Kingdom is the 2011 census workflow data [1]. Published by the Office of National Statistics under version 3.0 of the open government licence this data can be freely shared, adapted and republished for both commercial and non-commercial purposes [4]. We compare the BBC mobility data set to this census workflow data which only captures commuting patterns of adults to and from their usual place of work. We estimate - and systematically compare - human mobility models for the United Kingdom and impute national level origin-destination matrices for the total BBC data set and four coarse grained stratifications by age. We use these imputed commuting matrices to demonstrate how the risk of transmission between regions changes for each age-group compared to using aggregated data.

### 1.1 Human Mobility Models

So-called gravity models, where the rate of migration between spatially segregated populations is modelled as a function of the distance between locations and their relative population size, have proven to be exceptionally popular for epidemiological modelling [24, 45, 19, 42, 22, 12, 17, 25, 30, 33, 6]. Taking its name and form from the Newtonian law of gravitation, the theory actually has its origins in the social sciences and in particular transportation theory [20]. However, beyond the dependence on population size and distance there has been very little consistency in the definition of gravity models by different authors.

The most basic formulation of the gravity model has the convenient - but unrealistic - characteristic that the population flux between two locations only depends on the local characteristics of the two interacting populations. These foundational models take no account of the distribution of population - or connectivity - of the population between two points. The Radiation model [40] was developed to account for these effects explicitly by construction and in some human mobility data sets achieves similar – or better – predictive performance than gravity models despite having no free parameters to estimate. This parsimony comes at the expense of a lack of flexibility with the Radiation model being outperformed by classical gravity formulations when factors other than population density – such as the types and opportunities of travel – are more important. The extended radiation model [47] addressed this limitation by introducing a single scaling parameter *α* which can be interpreted as defining a characteristic length scale (*l*) for trips with a region.

There have been several attempts within the transport theory field to address these same issues within the gravity modelling framework - in particular the intervening opportunities model [41] and the competing destinations model [21]. We consider two variants of the intervening opportunities model – the Schneider formulation examined by [34] and Stoufer’s Rank Model as recently revisited in the context of modelling historical measles epidemics in England and Wales [2]. Finally, we consider the Impedance model [39]. Taking inspiration from the Ohmic law, the Impedance model was proposed as parameter free mobility model and compared to gravity and radiation laws in a model of the 2010 Haiti cholera epidemic [39].

Human mobility data sets typically capture a snapshot of individuals movement for which the total flux of individuals moving (or commuting) from a region is a fixed margin. For such data it is convenient to model the probability of moving given a particular home location separately from the choice of destination [40]. We therefore first compare the mobility rates (proportion of users that have different origin-destination locations) for the BBC data to census workflow data from the United Kingdom, then estimate constrained (singly or work constrained) variants of each mobility model [34].

All of these models have previously only been estimated from and tested against either aggregate mobility data or indirectly with respect to infectious disease case reports. The BBC mobility data set offers the first opportunity to compare and assess the predictive ability of these models for different groups of society.

## 2 Methods and Results

### 2.1 Data

There were two components to the BBC Pandemic study, one focused on the town of Haslemere [29], and another focused on the wider UK population [31]. Here we present mobility data from the UK national study. Upon starting the BBC Pandemic app users first entered their basic demographic information, including age, household size, gender and occupation. The app then recorded their location at a 1km grid scale across the UK at hourly intervals for a 24 hour period. At the end of this period, users provided key meta data including their gender, age, occupation, health today (self-assessed), number of people in household, maximum distance travelled (self-estimated) and mode of transport. Users were additionally asked to complete a detailed survey on the social contacts they made over the study period which are reported elsewhere [3].

Over 86,000 participants started the survey and filled out their profile. Participants with no encounter or location data were excluded, as were users whose location recordings were all outside the UK leaving 47,741 user with GPS trajectories. A small subset of users repeated the survey and provided multiple observation periods. To protect the anonymity of these users we only consider the location data collected during the first 24 hour period of observation for this study. As a further step to ensure anonymity of users we aggregated individual user trajectories to origin and destination locations at the mid-layer super output area (MSOA). As the mid-layer super output areas nest within the local area districts (LAD), we map the MSOA code to the corresponding LAD to define origin and destination locations at this higher spatial scale and calculate the final origin-destination flux matrices Ω_*ij*_ [8] that record the number of users with origin *i* and destination *j*.

We use the modal location to define users origin (home location) and considered two alternative definitions of destination based on the furthest extent and second (next) most frequent location from home. For the purposes of this study we focus on analysis on the ‘next’ (most frequent) origin-destination matrices as these are the most consistent conceptually with the census work flow data. (Full details of these definitions are given in supplemental information).

To provide some context on changing patterns of mobility over the working week we tabulate the start time of each individuals observation period aggregated by day of week (Table 3). The majority of users started the app on a weekday (37,322). The highest single day of sign-up was on the Thursday that the associated BBC Pandemic documentary was originally broadcast (22nd March 2018). 2,981 users signed up during the hour of broadcast alone, with a total of 7,796 users starting the app on that day. The next highest day of sign-up (with 4,967 new users) was also a Thursday (28th September 2017) corresponding to the second day of a social media campaign run by the production company to promote the app.

Given the relative sparseness of the data set at the MSOA level, we focus on analysing the patterns of mobility at the LAD level. Raw data, estimated models and imputed flux matrices for both the next and furthest extent definitions are available in a public repository along with code developed for this manuscript (https://github.com/BBCPandemic/BBCMobility).

Of the 43,291 users in the BBC mobility data set (henceforth the Total BBC data), 25,114 had inferred home and next most frequent locations within the same LAD. We define the complementary set of 18,177 users with origin and destination locations in different LADs as ‘movers’ (Tables 1,2, Figure 1) rather than commuters to acknowledge that the displacements in the BBC mobility set capture a wider range of human mobility than the strictly commuting flows measured by census work flow data.

**Table 1:** Number of BBC users by member nation of the UK and number of records from census workflow data. Users are the total individuals with a paired origin-destination. Movers are the number of individuals whose destination location is different from their home location. We estimate the average per capita probability of moving for each data set by the ratio of these two numbers (presented to 3 s.f.).

| **Data Set** | **Users** | **Movers** | **P(move) per capita per day** |
| --- | --- | --- | --- |
| Total | 43,291 | 18,177 | 0.420 |
| BBC England | 37,852 | 16,613 | 0.438 |
| BBC Wales | 1,699 | 535 | 0.315 |
| BBC Scotland | 3,092 | 867 | 0.280 |
| BBC NI | 548 | 162 | 0.296 |
| Census (E) | 23,139,206 | 9,953,850 | 0.430 |
| Census (W) | 1,263,873 | 383,303 | 0.303 |
| Census (NI) | 724,873 | 199,052 | 0.275 |
| Census (Scotland) | 2,242,725 | 619,372 | 0.276 |

**Table 2:** Number of BBC users by age and employment categories. Users are the total individuals with a paired home and work location. Movers are the number of individuals whose destination location is different from their home location. We estimate the average per capita probability of moving for each data set by the ratio of these two numbers (presented to 3 s.f.). Note that Under 18 is a category in both the age-stratified and employment data sets but is presented twice to emphasise that these are two distinct (not nested) stratifications of the total BBC mobility data set. Age ranges are open on the lower limit and closed on the upper: (0, 18], (18, 30], (30, 60], (60, 100].

| **Data Set** | **Users** | **Movers** | **P(move) per capita per day** |
| --- | --- | --- | --- |
| BBC Under 18 | 2,955 | 724 | 0.245 |
| BBC 18-30 | 9,611 | 4,015 | 0.418 |
| BBC 30-60 | 26,009 | 11,948 | 0.460 |
| BBC 60+ | 4,716 | 1,490 | 0.316 |
| BBC Under 18 | 2,955 | 724 | 0.245 |
| BBC Education | 3,522 | 1,101 | 0.314 |
| BBC Employed | 30,500 | 14,710 | 0.482 |
| BBC NEET | 6,325 | 1,42 | 0.260 |

**Figure 1:**
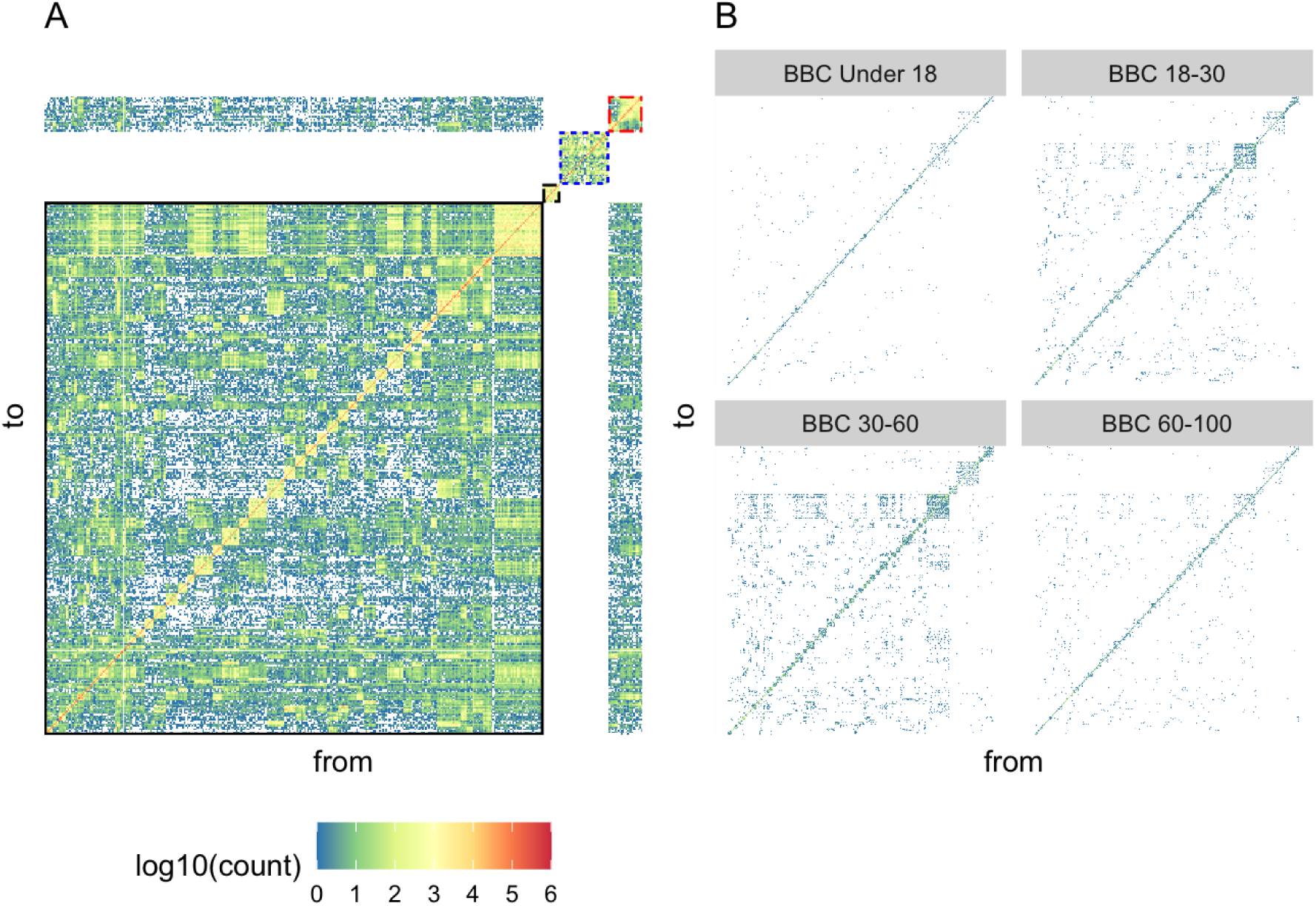
Comparison of raw population flux from the 2011 census and BBC Data Set. (A) Heat map of the frequency of reported work flows between the 391 local administrative districts (LADs) of the United Kingdom. Colour scale (shared between panels) is logarithmic (log10(count) in each cell), with white indicating that no flux was recorded for a given pair of locations. The 2011 census collected self-reported home (from) and work (to) locations which are collated and published separately for England & Wales, Northern Ireland and Scotland. The national work flows therefore take a block diagonal in the flux matrix, with white gaps representing the missing sub-national work flows. From bottom left to top right, the first block matrix is the workflows within England (solid black outline), Northern Ireland (dashed black outline), Scotland (dotted blue outline) and Wales (dash-dot red outline). The outer off-diagonal elements represent the intra-national work flows between England and Wales. (B) Heat maps of the BBC mobility flux data sets stratified by age category (top left - bottom right): Under the age of 18, greater than 18 and less than 30 (BBC 18-30), greater than 30 and less than 60 (BBC 30-60) and greater than 60 (BBC 60+).

#### 2.1.1 Modeling the origin-destination flux matrix

Flux patterns can be decoupled into two model components: the probability that someone moves between locations, and the location that someone goes to given that they do move. The origin-destination flux matrix Ω_*ij*_ can be correspondingly decomposed. The diagonal elements represent users who stayed in the same location and the off-diagonal elements correspond to the ‘movers’. Thus, the flux matrix of movers alone is given by the origin-destination flux matrix with the diagonal elements set to zero:

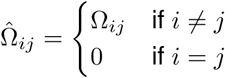

The total eflux from location *i* is given by the sum over all potential destinations 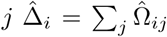 while the total number of users with origin *i* (*i*.*e*. including the non-movers) is Δ_*i*_ = _*j*_ Ω_*ij*_. The proportion of movers for location *i* is given by 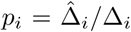. We define the conditional movement matrix:

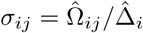

as the proportion of movers from *i* who choose destination *j*. It can then be seen that *σ* satisfies the normalisation Σ_*j*_ *σ*_*ij*_ = 1 and by definition *σ*_*ii*_ = 0. The origin-destination flux matrix can thus be decomposed in terms of the conditional movement matrix *σ*_*ij*_ and probability of movement *p*_*i*_:

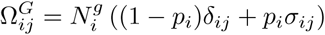

where *δ*_*ij*_ is the alternating Kronecker delta symbol.

#### 2.1.2 Probability of moving

The data are now summarised in two separate components: the proportion of the population that move (*p*_*i*_) and where the movers go (*σ*_*ij*_). We first we consider the first part of this: the probability of movement, comparing the BBC and census datasets, and also explore how probability of movement varies by age and employment status.

Census outputs are prepared and published separately for Scotland, Northern Ireland and England & Wales at different levels of spatial aggregation. The Northern Ireland Statistics and Research Agency excludes all responses with work locations outside of Northern Ireland. The Office of National Statistics and Scotland’s Census publish aggregate numbers for the total work flows to each member nation, but not the sub-national location. For consistency, and to make comparisons between the member nations of the United Kingdom, we separate the combined England and Wales data set and treat the four resulting public census outputs as separate data sets for the purposes of model estimation and comparison (Table 1). Sub-national movement rates are comparable between the census workflow and BBC mobility data sets (to 1 significant figure).

The meta-data collected by the BBC pandemic app allows us to further stratify the Total BBC data set by age and employment categories. For age we consider four coarse grained categories – under the age of 18 (BBC Under 18), over 18 and under 30 (BBC 18-30), over 30 and under 60 (BBC 30-60) and over the age of 60 (BBC 60+). With respect to employment status we reuse the under 18 category (BBC Under 18), and define three alternative subsets for analysis corresponding to the group of users over the age of 18 and in education (BBC Education), over the age of 18 and in employment (BBC Employment) and over the age of 18 and not in employment, education or training (BBC NEET). The per-capita mobility rate for these strata of the BBC mobility data set vary from a minimum of 0.245 for BBC Under 18 to a maximum of 0.460 for BBC 30-60 (Table 2).

#### 2.1.3 Geographic variation

The proportion of movement *p*_*i*_ also varies considerably between regions (Figure 2). There is no clear relationship with the size of the resident population and only a weak association between the area of LADS and *p*_*i*_. However, there is a clear linear relationship between *p*_*i*_ from the BBC data and the corresponding probability of movement from the census data, with an *R*^2^ = 0.71, suggesting that both instruments are measuring a common source of variability in mobility between regions.

**Figure 2:**
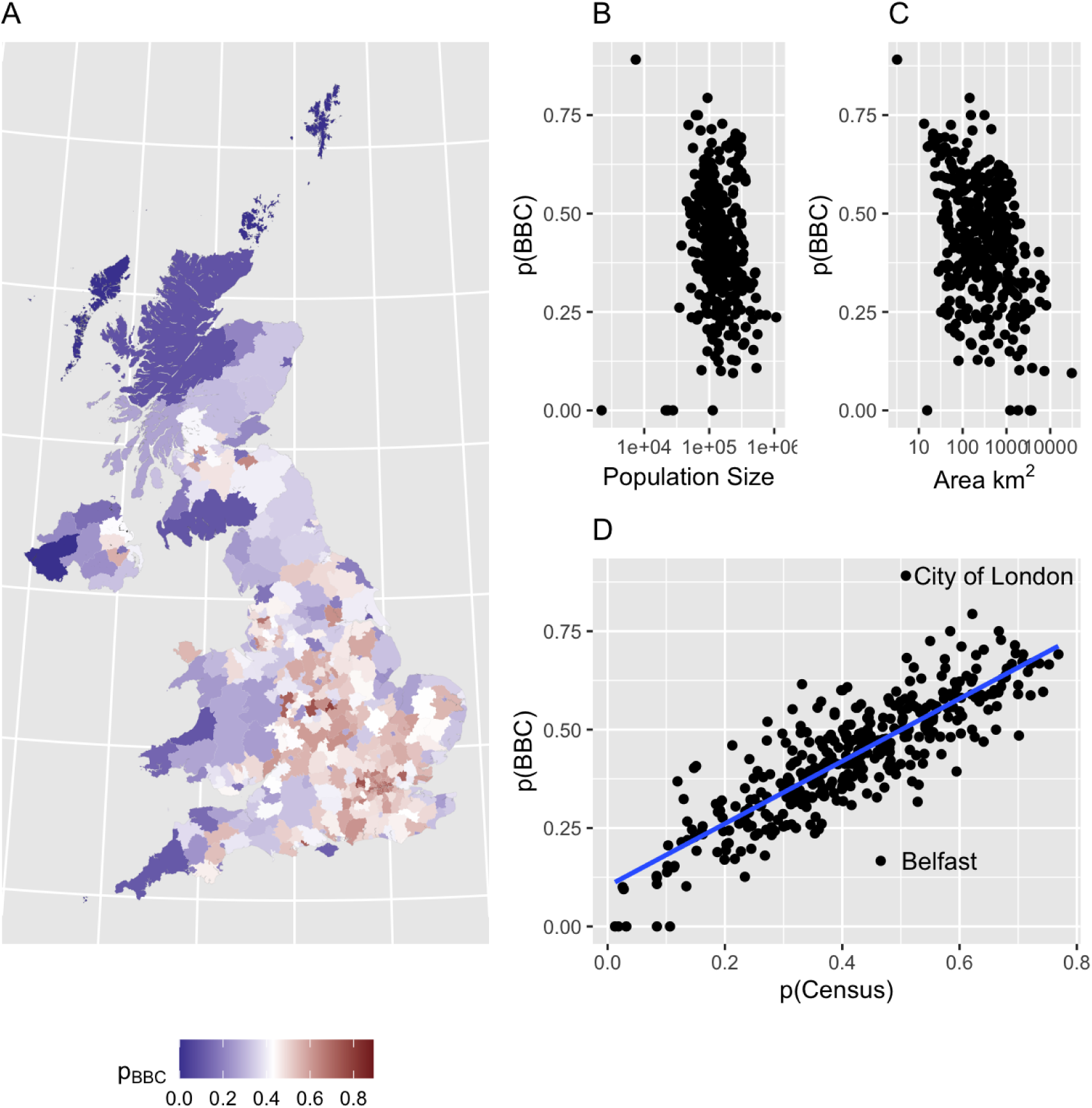
Geographic variation in probability of movement by LAD. (A) Heat map of the probability of movement (*p*(*BBC*)), defined as fraction of users with differing origin and destination LADS, for the 391 local administrative districts (LADs) of the United Kingdom. Colour scale is centred on the the national average (*p*(*BBC*) =0.420, white). *p*(*BBC*) has no relationship with the resident population size (B), but has a weak inverse relationship with the area of the LAD (C). The geographic variation in *p*(*BBC*) does have a clear linear relationship (D) with the corresponding fraction of movers from the UK census workflow data (*p*(*Census*)). A simple linear regression (blue line) has an R-squared = 0.71, with *p*(*BBC*) = 0.1 + 0.79*p*(*Census*)

With a median of 81 users per LAD (range 2-948) the raw BBC data is too sparse to estimate the geographic variation in population flux from each LAD for each stratification of interest (i.e. age or employment status). To address this limitation and be able to impute movement rates for each age and employment group we model the per location probability of moving using a generalised linear model (with logit link) and random intercept term for each LAD (*i*):

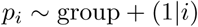

where group is a categorical variable that adjusts the background rate of movement for each level of the age or employment strata. We therefore assume that the difference in mobility between groups is constant across the UK, proportionally adjusting the regional mobility captured by the random intercept term. Random effects models were estimated using the lme4 package [11] in the R statistical language [38].

Model fit was assessed graphically by plotting the predicted probability of movement against the observed data (Supplemental Figures 9,10). This basic check illustrates the model is performing as intended, fitting closely to the 18-30 and employed categories which have the largest sample size and pulling the imputed movement rate for under-sampled LADS and categories (Under 18, NEET) towards the group and local (LAD) averages.

### 2.2 Mobility Models

Here we now focus on the model component of where people go, conditional that they do move, in essence modelling the conditional movement matrix *σ*_*ij*_ as defined above.

#### 2.2.1 Gravity Model

The classical gravity formulation assumes that the flux between two locations depends on the product of the size of the donor (*N*_*i*_) and destination (*N*_*j*_) populations (with scaling parameters *τ*_2_, *τ*_1_) divided by a function of the relative distance (*r*_*ij*_) between them:

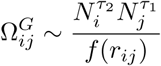

However, this general form is arguably *too* flexible, and in particular is unbounded with no upper limit on the predicted number of commuters from the donor patch. We can normalise the gravity model by defining a vector of normalisation constants 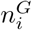 for each donor patch *i* by summing over the set of recipient patches *j* (for all (*j* ≠ *i*):

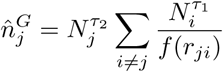

forming a so called singly constrained gravity model. Recasting the model in terms of the conditional movement matrix *σ*_*ij*_, the donor term 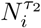 cancels out and is redundant in this formulation. We therefore define our basic gravity law model for *i* ≠ *j* as:

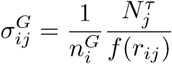

with *σ*_*ii*_ = 0 by definition and:

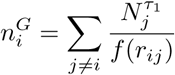

We consider gravity-type models with three different distance scaling functions, power-law 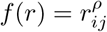, exponential *f* (*r*) = *e*^*rij/ρ*^ and offset 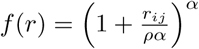.

#### 2.2.2 Competing Destinations Model

Following the same reasoning, the competing destinations model [21] can be defined for our purposes for *i* ≠*j* as:

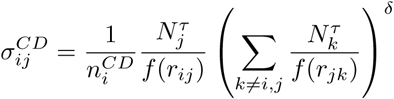

where:

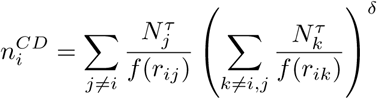

The parameter *δ* adjusts for the effect that other locations – the competing destinations – have on the flux. For a negative value of *δ* the effect of competing destinations is to reduce the flux to a particular location, whereas for a positive *δ* the flux between the two locations is enhanced by the presence of alternative destinations. When *δ* = 1 the competing destinations term vanishes and we recover the classical gravity model. As the gravity model is nested within competing destinations we do not fit these simpler gravity model formulations separately. We therefore compare three gravity type models: competing destinations with power law distance function (CDP), competing destinations with exponential distance function (CDE) and competing destinations with offset distance function (CDO).

#### 2.2.3 Extended Radiation Model

The radiation model has no free parameters to estimate and is normalised by construction:

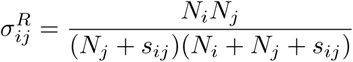

where *s*_*ij*_ is the total population in a circle of radius *r*_*ij*_ centred at *i* and excluding *N*_*i*_ and *N*_*j*_ themselves [40]. The extended radiation model (ERad) introduces a single scaling parameter *α* [47].

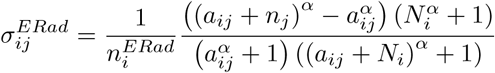

where

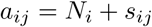

Yang et al. [47] proposed that *α* should vary between between regions with different characteristic length scales (*l* defined as mean length between regions being modelled) such that:

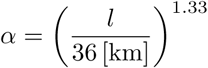

For the UK local authorities, McNeill et al. [36] calculated *l* = 19km and thus we would expect *α* = 0.43. Here we estimate *α* as a free parameter to compare with this predicted scaling law.

#### 2.2.4 Intervening Opportunities Model

Schneider’s intervening opportunities (IO) model [34] depends on the same matrix *s*_*ij*_ as the radiation model and is defined for *i* ≠ *j* as:

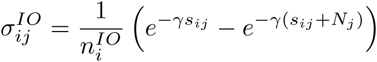

where

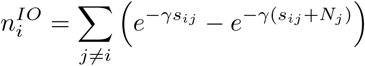

#### 2.2.5 Stoufer’s Rank Model

Stoufer’s (Sto) Rank model [2] also depends on the same matrix *s*_*ij*_ as the radiation model and can be defined as:

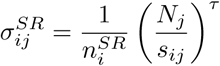

where

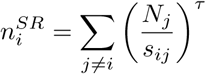

#### 2.2.6 Impedance Model

The impedance (Imp) model [39] is defined as:

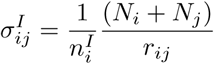

where

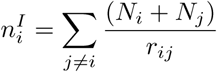

In common with the radiation model this mobility model has no free parameters to estimate.

### 2.3 Inferential framework and model comparison

Each row 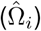 of the mover flux matrix 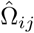 could be considered as a multinomial sample [42] with 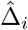 trials and probability vector *σ*_*i*_ equal to the corresponding row of the mobility matrix *σ*_*ij*_:

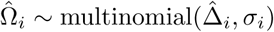

However, for a multinomial likelihood the variance of observations scales linearly with the number of trials. As the efflux 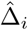 scales with local population size, a multinomial likelihood for these data will be dominated by contributions from large urban centres potentially introducing systematic biases and not allowing for the possibility of over-dispersion in the sampled flux. The expected rate of flux for a given mobility matrix *σ*_*i*_ is:

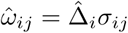

From which we can construct a negative binomial likelihood and explicitly allow the variance to scale with (origin) population size:

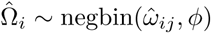

Note that we use the ecological parameterisation of the negative binomial specified by the mean and shape parameter *ϕ*) and the variance of the flux leaving patch *i* is thus 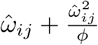..

We estimate posterior distributions for each model using Hamiltonian MCMC (as implemented by Stan [16] http://mc-stan.org/). To assess model fit and provide a basis for model selection we use approximate leave-one-out cross-validation [43, 44]. For numerical stability we restrict the range of parameters such that 0 *< τ, ϕ <* 5, 0 *< α <* 1, − 5 *< δ <* 5 and 0 *< γ <* 10^*−*4^. We restrict *ρ >* 0 for the offset and exponential competing destinations models (CDO, CDE), with the further restriction that 0 *< ρ <* 5 for the power law scaling (CDP). We choose Cauchy (0,5) prior distributions for all parameters. All further analyses were carried out in R [38].

### 2.4 Model checking and cross-validation

For each combination of model and data set, 4 Hamiltonian MCMC chains were run for the default 2,000 iterations, unless a greater number were required to pass diagnostic checks. Chains were well mixed for all combinations of models and data sets and passed standard convergence diagnostics. As a further predictive check of the model – and to provide a basis for model comparison and selection – we carried out approximate leave-one-out cross validation (LOO) [43].

This method uses Pareto smoothed importance sampling (PSIS-LOO) [44] to estimate the expected log pointwise predictive density: 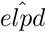 which measures the predictive accuracy of the model when a single observation is dropped out. The difference 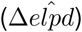 between 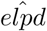 for alternative models fitted to the same data provides a measure of their relative predictive accuracy. The standard error on the difference gives a measure of uncertainty with standard errors comparable to the magnitude of the difference suggesting the relative predictive accuracy of the two models is indistinguishable.

The estimated (Pareto) shape parameters 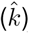 for the predicted distribution of 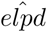 can be used to judge the reliability of the estimate of 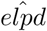 for each data point. The estimate of 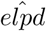 is considered reliable for 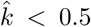, performance may still be reliable for values of 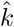 up to 0.7. Values of 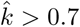 suggest that the data points are highly influential to the estimated posterior and potentially introducing bias.

Models estimated from the full BBC mobility and Census data sets have five locations with 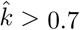 and two with 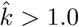 suggesting these locations are highly influential to the estimated posterior distribution and potentially introducing bias. Most of these issues can be traced to the uniquely large number of commuters to two specific districts within London: namely the City of London and Westminster. The City of London and Westminster (and to a lesser extent other boroughs of London) attract anomalously large numbers of commuters for their size. This can be visualised by plotting the numbers of commuters in (influx) against leaving (efflux), (Supplementary figure 11). For the majority of LADs this relationship is symmetric with both the influx and efflux of commuters approximately proportional to the resident population size. However the City of London and Westminster have orders of magnitude higher commuters in than residents who commute out. This extreme lack of fit of gravity type models results in a systematic bias to parameter estimates as illustrated by comparing the posterior estimates between the full set of 326 English LADs and a data set dropping the City of London and Westminster (324 English Lads, Supplementary figure 12).

The Highland LAD in Scotland has a similar impact on parameter estimates from the BBC mobility data set (due in this case to the small flux of users to this location). Removing these three locations, then all of the models estimated from the BBC mobility data have 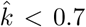. However, even after these steps between 1 to 9 LADS have 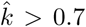. for the gravity type CDO, CDE and CDP models estimated from census data. Given the greater overall predictive performance of these models on the BBC mobility data set we do not wish to simply set aside these models.

Although the specific problematic LADS vary between models, the value of 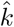 increases directly with the size of the resident population. Gravity models are notoriously sensitive to contributions from high population density locations. This, and the disparity in sample sizes between the BBC mobility and census data sets, was our motivation for using a negative binomial likelihood where the shape parameter *ϕ* controls the variance to mean scaling relationship. For a mean flux *y*, the variance of the negative binomial likelihood is inversely related to the shape parameter 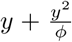. Reducing the value of *ϕ* reduces the influence of larger populations to the likelihood, hence we can carry out a sensitivity analysis to the influence of the outlier values by fixing the value of *ϕ* and reducing it until the pareto shape parameter for all of the observations *<* 0.7. This was achieved for a fixed shape parameter of *ϕ* = 0.1. The models estimated from census data with a fixed value of *ϕ* reduced the absolute values of the likelihood but did not change the rank ordering of models (described below). Although the point estimates of the gravity parameters do change systematically with *ϕ*, they still have overlapping credible intervals in the range between the estimated value and fixed value (0.1) where the census models pass all diagnostic checks (13). For the purposes of comparison to the BBC mobility data set we present the models with estimated value of *ϕ*, which have a greater overall predictive performance (as measured independently via the common part of commuters index described in the next section).

### 2.5 Model Selection and Predictive Performance

The rank order of mobility models based on 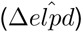 is identical for the UK level stratifications of the BBC mobility data set (Tables 5, 6, 7) – with the competing destinations (CDO) model favoured above the other models for all data sets. The ranking of the second and third ranked models varies between member countries of the UK, but the differences are small compared to the standard deviation implying there is little to choose between the predictive ability of the three gravity formulations for these data. The English census data is the only data set where an alternative model – the Extended Radiation model is preferred.

**Table 3:** Number of users by weekday of start date.

| Weekday | Users |
| --- | --- |
| Monday | 3,340 |
| Tuesday | 3,476 |
| Wednesday | 6,945 |
| Thursday | 16,094 |
| Friday | 7,467 |
| Saturday | 3,168 |
| Sunday | 2,801 |

**Table 4:** Mobility model comparison for the sub-national Census workflow data sets. Mobility models are ranked by their predictive accuracy as measured by the difference 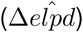 in the expected log pointwise predictive density 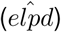. The competing destinations model, with offset distance kernel, is favoured for the census workflow data from Wales (W), Scotland (S) and Northern Ireland (NI). For the English census data, the Extended radiation model is ranked first. The difference in predictive accuracy between the top ranked models is greater than the standard error for the English census data, but of a comparable magnitude for the relatively smaller Scottish, Welsh and Northern Irish data sets. The differences between the next ranked models are much smaller than with respect to the top ranked model.

| E Census |  | W Census |  | S Census |  | NI Census |  |
| --- | --- | --- | --- | --- | --- | --- | --- |
| Model | $\Delta elpd$ (s.d.) | Model | $\Delta elpd$ (s.d.) | Model | $\Delta elpd$ (s.d.) | Model | $\Delta elpd$ (s.d.) |
| **ERad** | 0 (0) | **CDO** | 0 (0) | **CDO** | 0 (0) | **CDO** | 0 (0) |
| CDO | -11500 (1100) | CDP | -23.2 (7.8) | CDP | -16.4 (10) | CDE | -4.18 (3.2) |
| CDP | -12300 (1100) | CDE | -82.5 (17) | IO | -239 (51) | CDP | -32.6 (9.4) |
| IO | -35400 (640) | IO | -278 (32) | ERad | -256 (45) | IO | -50.9 (8.2) |
| CDE | -41400 (850) | ERad | -302 (26) | CDE | -351 (37) | ERad | -65 (7.2) |
| Imp | -60900 (880) | Imp | -392 (31) | Imp | -465 (44) | Imp | -78.9 (5.1) |
| Sto | -79300 (960) | Sto | -486 (33) | Sto | -644 (44) | Sto | -98.8 (6.3) |

**Table 5:** Mobility model comparison for the sub-national BBC Total mobility data sets. Mobility models are ranked by their predictive accuracy as measured by the difference 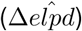 in the expected log pointwise predictive density 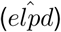. The competing destinations model, with offset distance kernel, is favoured for all four subsets of the BBC Total mobility data from England (E), Wales (W), Scotland (S) and Northern Ireland (NI). The difference in predictive accuracy between the top ranked models is greater than the standard error for the English census data, but of a comparable magnitude for the relatively smaller Scottish, Welsh and Northern Irish data sets. As with the census data sets the differences between the next ranked models are much smaller than with respect to the top ranked model.

| BBC Total (E) |  | BBC Total (W) |  | BBC Total (S) |  | BBC Total (NI) |  |
| --- | --- | --- | --- | --- | --- | --- | --- |
| Model | $\Delta elpd$ (s.d.) | Model | $\Delta elpd$ (s.d.) | Model | $\Delta elpd$ (s.d.) | Model | $\Delta elpd$ (s.d.) |
| **CDO** | 0 (0) | **CDO** | 0 (0) | **CDO** | 0 (0) | **CDO** | 0 (0) |
| CDE | -287 (41) | CDE | -3.64 (2.7) | CDE | -12.5 (8.6) | CDP | -0.562 (2.7) |
| ERad | -319 (76) | CDP | -9.56 (4.4) | CDP | -14.3 (10) | CDO | -1.9 (2.9) |
| CDP | -2270 (110) | IO | -46.6 (9.2) | IO | -34.1 (16) | ERad | -4.9 (2.1) |
| IO | -3230 (110) | ERad | -51.2 (8.2) | ERad | -45.6 (19) | CDE | -4.96 (3.8) |
| Imp | -6330 (160) | Imp | -80.1 (11) | Imp | -123 (23) | Imp | -27 (5) |
| Stoufer | -10400 (220) | Stoufer | -151 (14) | Stoufer | -220 (27) | Stoufer | -42.2 (6.3) |

**Table 6:** Mobility model comparison for the BBC Mobility data for the UK stratified by age category. Mobility models are ranked by their predictive accuracy as measured by the difference 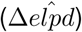 in the expected log pointwise predictive density 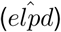. The competing destinations model, with offset distance kernel, is favoured for all data sets. In contrast to the sub-national data sets the ranking of models estimated from the BBC data sets are consistent across age groups and the difference between the predictive accuracy of the top ranked model is greater than the standard deviation of the difference. The differences between the second ranked models, in this case the exponential distance kernel and Extended Radiation models, are once again much smaller than the difference with respect to the favoured model (CDO).

| BBC Under 18 |  | BBC 18-30 | BBC 30-60 | BBC 60+ |
| --- | --- | --- | --- | --- |
| Model | $\Delta elpd$ (s.d.) | $\Delta elpd$ (s.d.) | $\Delta elpd$ (s.d.) | $\Delta elpd$ (s.d.) |
| **CDO** | 0 (0) | 0 (0) | 0 (0) | 0 (0) |
| CDE | -21.1 (7.1) | -95.8 (18) | -247 (35) | -30.3 (9.4) |
| ERad | -67 (18) | -182 (38) | -336 (67) | -115 (21) |
| CDP | -154 (28) | -843 (89) | -2100 (130) | -397 (41) |
| IO | -395 (48) | -1350 (82) | -3130 (110) | -749 (48) |
| Imp | -795 (42) | -2450 (94) | -5710 (150) | -1080 (52) |
| Stoufer | -1340 (66) | -4850 (170) | -10100 (220) | -2330 (88) |

**Table 7:** Mobility model comparison for the BBC Mobility data for the UK stratified by employment category. Mobility models are ranked by their predictive accuracy as measured by the difference 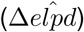 in the expected log pointwise predictive density 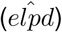. The competing destinations model, with offset distance kernel, is favoured for all data sets. In contrast to the sub-national data sets the ranking of models estimated from the BBC data sets are consistent across employment groups and the difference between the predictive accuracy of the top ranked model is greater than the standard deviation of the difference. The differences between the second ranked models, in this case the exponential distance kernel and Extended Radiation models, are once again much smaller than the difference with respect to the favoured model (CDO).

|  | BBC Total | BBC Under 18 | BBC Education | BBC Employed | BBC NEET |
| --- | --- | --- | --- | --- | --- |
| Model | $\Delta \hat{elpd}$ (s.d.) | $\Delta \hat{elpd}$ (s.d.) | $\Delta \hat{elpd}$ (s.d.) | $\Delta \hat{elpd}$ (s.d.) | $\Delta \hat{elpd}$ (s.d.) |
| **CDO** | 0 (0) | 0 (0) | 0 (0) | 0 (0) | 0 (0) |
| CDE | -299 (43) | -21.1 (7.1) | -19.9 (6.4) | -297 (40) | -27.9 (9.6) |
| ERad | -438 (81) | -67 (18) | -33.7 (20) | -449 (73) | -98.7 (26) |
| CDP | -2950 (150) | -154 (28) | -298 (38) | -2410 (140) | -525 (52) |
| IO | -4130 (120) | -395 (48) | -530 (44) | -3520 (120) | -872 (52) |
| Imp | -7580 (170) | -795 (42) | -803 (44) | -6600 (160) | -1230 (57) |
| Stoufer | -12500 (250) | -1340 (66) | -1660 (80) | -11300 (240) | -2560 (91) |

As a final posterior predictive check of the estimated models we consider the Common Part of Commuters (CPC) index introduced by [47] which can be defined as:

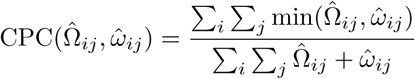

where 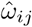 is a posterior predictive flux matrix from a mobility model fitted to the empirical flux matrix 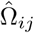 and the sum is over all donor (*j*) and recipient (*i*) patches within the meta-population. A value of 1 is calculated when there is perfect agreement between the two matrices, with 0 when there is no agreement. Figure 3 presents the posterior predictive distributions for the CPC for each combination of mobility model and data set. This measure is in broad agreement with the ranking achieved through LOO cross-validation - with the competing destinations model favoured in the majority of cases, but once again with relatively little difference in predictive performance between the top three models.

**Figure 3:**
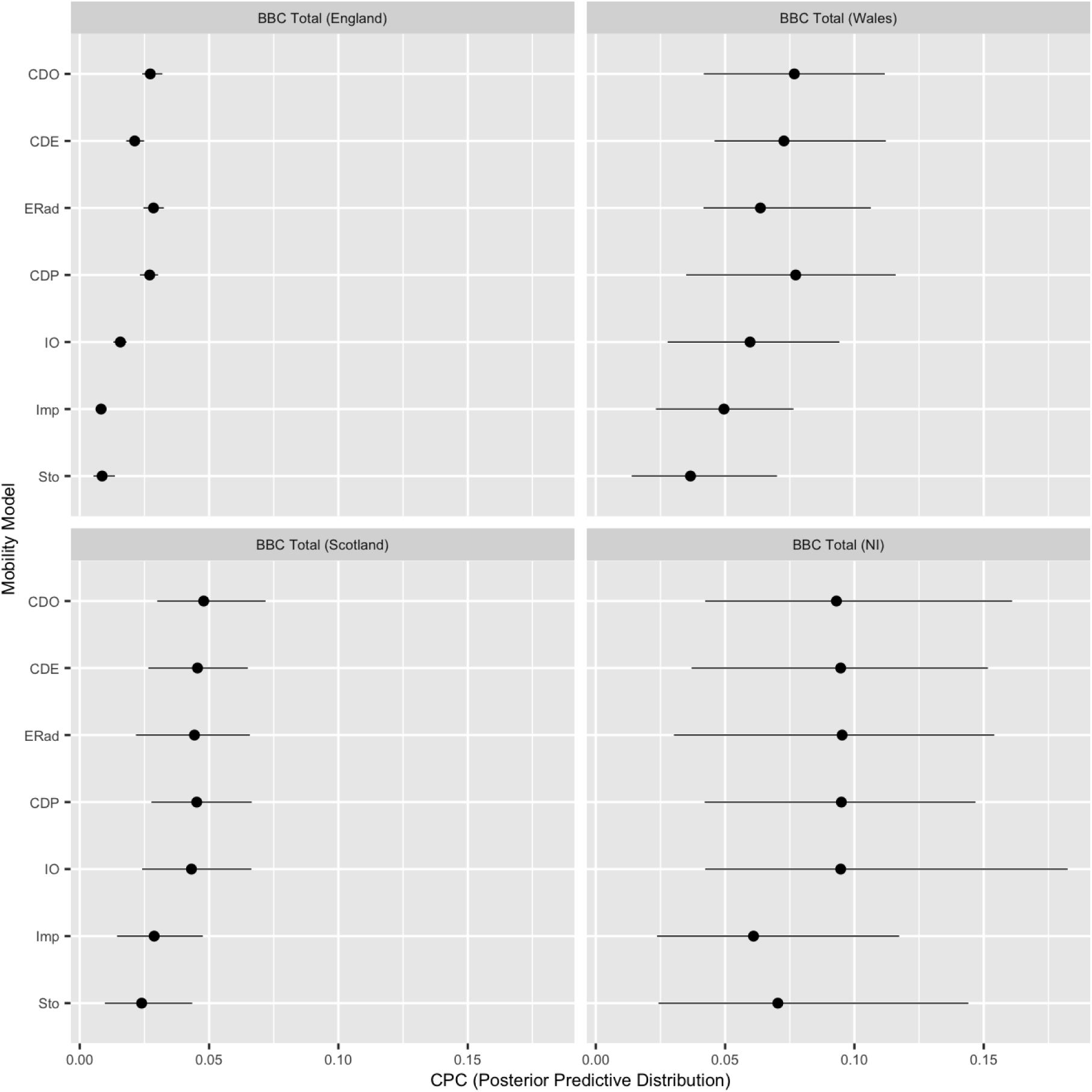
Posterior predictive distributions for the Common Part of Commuters (CPC) index. The Common Part of Commuters (CPC) index measures the agreement between the posterior predicted flux between each location and the empirical flux used to estimate the model with 1 indicating perfect agreement and 0 no agreement. Performance of all seven candidate mobility models (Competing Destinations - CDO, CDP, CDE, Extended Radiation - ERad, Stoufer’s Rank Model - Sto, Impedance - Imp) is compared for (top left panel through to bottom right) the (sub-sampled) census data sets (England, Wales, Northern Ireland - NI and Scotland, sub-national subsets of the BBC Total data set. Note that the predicted flux for the Impedance models depends on no free parameters, so for these models the average CPC is a function only of the topological distribution of local administrative districts for the United Kingdom and the member countries (census data). The variance of the predictive distribution for CPC for the Impedance models does vary between data sets through the estimated shape parameter. The rank ordering is consistent with the results of the LOO analysis, as are the relatively small predictive differences between the three gravity type models and the Extended radiation model.

### 2.6 Impact of age and employment status on mobility patterns

We first compare the gravity parameters for the BBC Total data set estimated from each of the member nations of the UK and compare with estimates from the census workflow data (Figure 4). The pooled UK estimates are most consistent with Census estimates from England (which has the largest population and number of LADs) but there are systematic differences in estimates from England and the smaller member nations. Containing only 10 LADs the Northern Ireland data set is clearly to small to support inference of this model, with huge uncertainty in the results should not be interpreted and are presented only for completeness. The Scottish and Welsh data sets demonstrate an increased importance of population size (larger *τ* value) and a longer spatial scale (*ρ, α* parameters) compared to estimates from England and the pooled UK data.

**Figure 4:**
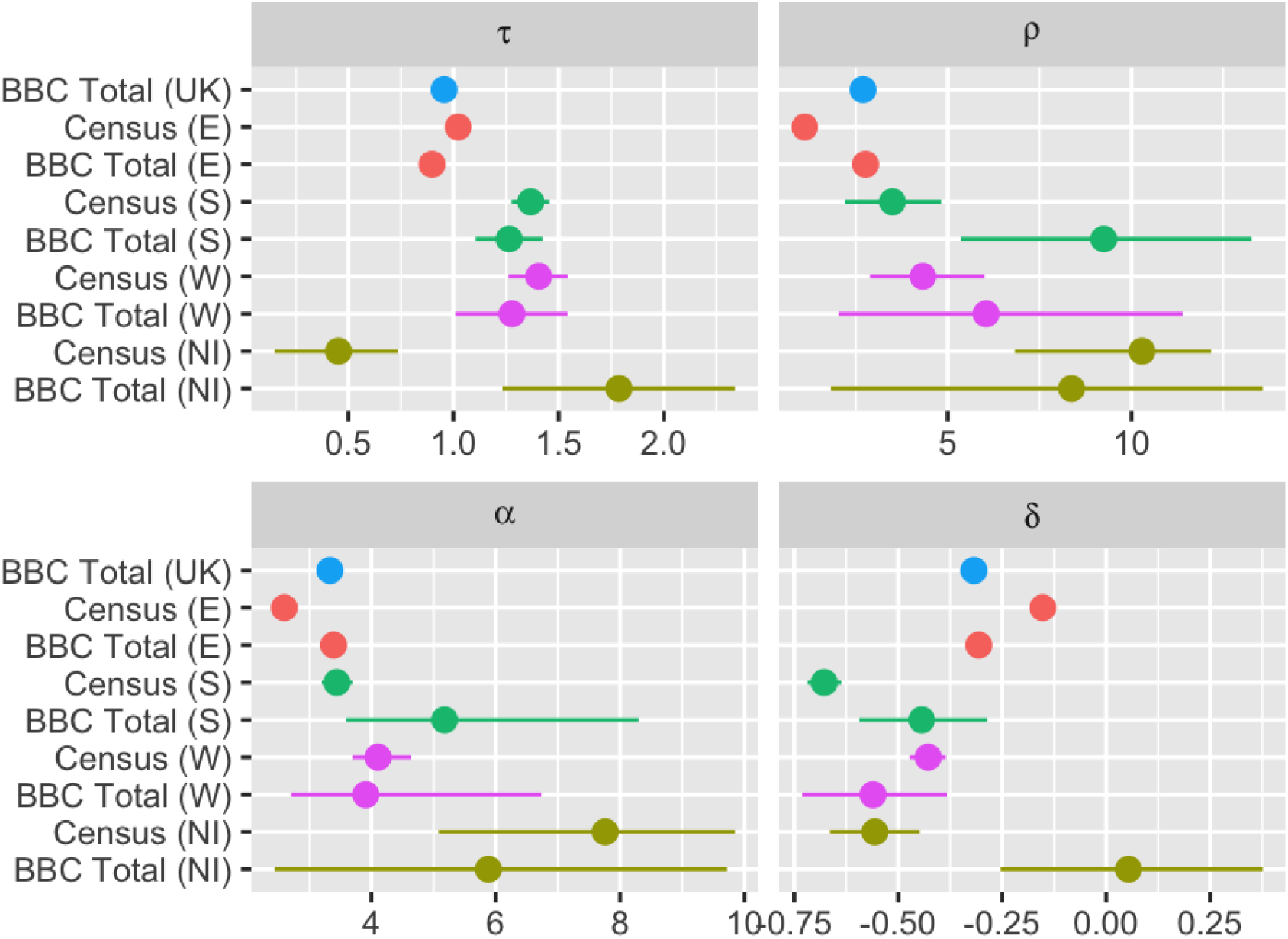
Sub-national posterior estimates for the competing destinations (CDO) model. Posterior distributions for the population density (*τ*), distance (*ρ, α*) and competing destinations (*δ*) scaling parameters from the (sub-sampled) Census data from England, Wales, Scotland and Northern Ireland and the corresponding subsets of the BBC mobility data. At the aggregate level parameter estimates are consistent between the BBC Total and Census data sets reflecting systematic differences in mobility patterns between England, Scotland and Wales. Northern Ireland estimates presented for completeness – note the considerably larger uncertainty resulting from the small size of this data set.

Given this sub-national variation it would be ideal to compare the mobility patterns of by age and employment categories of the BBC mobility data set at this level. However, given the sample size we must restrict comparison to models estimated at the national (UK) level. There is close correspondence between the age and employment groups suggesting that the 18-30, 30-60 and 60+ age groups behave largely like the education, employed and NEET categories.

Once again given that the vast majority of users were are in the 30-60 or employed categories, the parameters estimated from the BBC Total data set are statistically indistinguishable from those from the subset that were employed with overlapping posterior distributions (Figure 5). There are systematic differences in the estimated gravity scaling parameters for Under 18s, those in Education and NEETS. Under 18s have a faster decay in mobility with distance than other groups (larger *α*) and experience the strongest impact of competing destinations. Population size is a more important predictor for those over the age of 18 and in full time education (higher estimated value of *τ*) - which may reflect the location of universities, colleges and other higher education institutions within urban centres. By comparison NEET’s range further (smaller *α*, but population density is less important in modulating their decisions (smaller *τ*).

**Figure 5:**
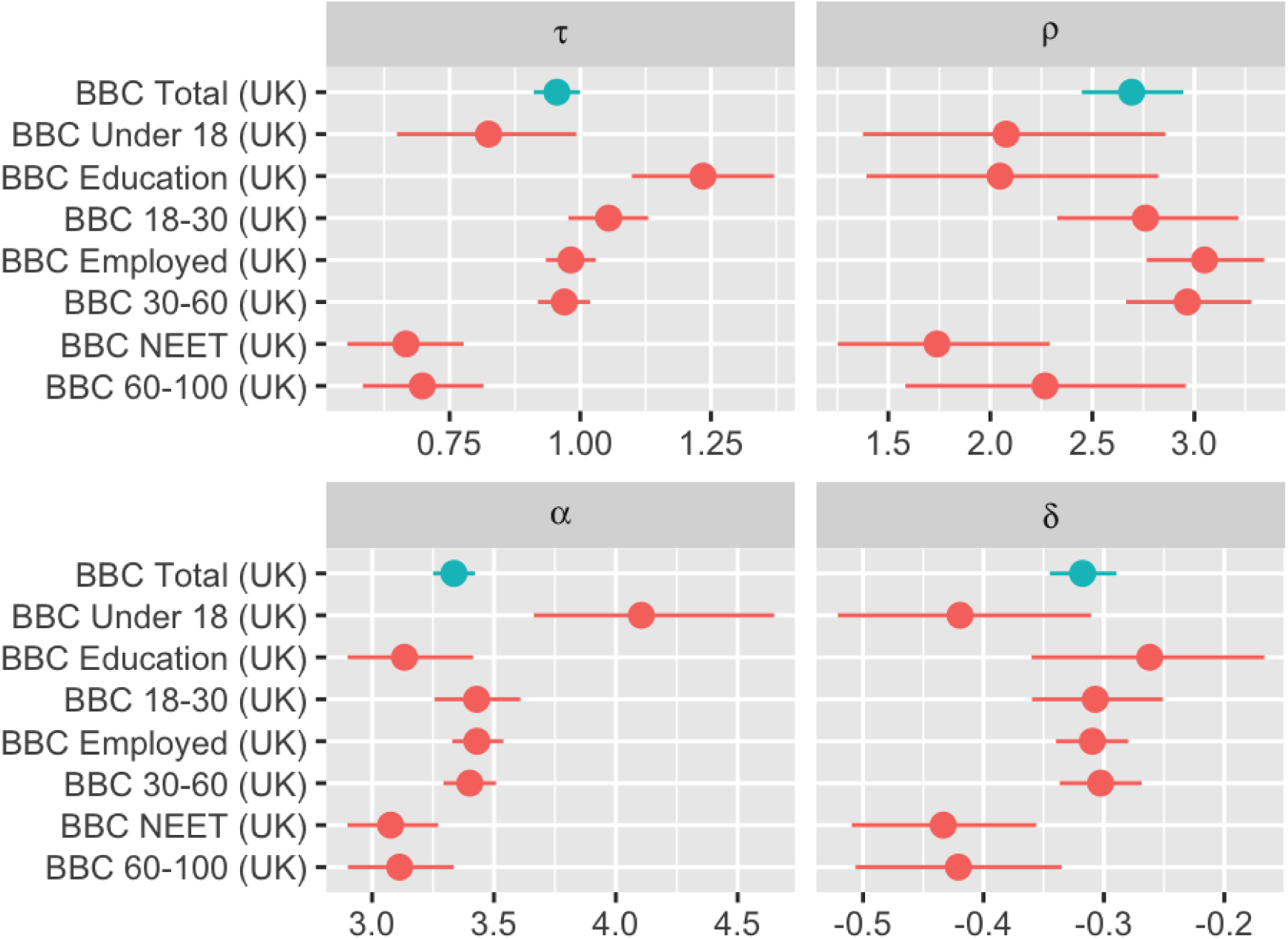
Posterior estimates for the competing destinations (CDO) model by age and employment groups. Posterior distributions for the population density (*τ*), distance (*ρ, α*) and competing destinations (*δ*) scaling parameters estimated from the full UK BBC mobility data set, with comparison between models estimated to the stratified data sets with respect to age and employment groups.

To facilitate the use of the BBC mobility data in simulation studies, we use our estimated models for the probability of movement 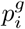 and choice of destination 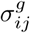 to impute UK population flux matrices for each level of the BBC mobility data set (Figure 6):

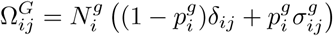

where 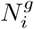 is the total population in group *g* resident in patch *i*. These imputed matrices are calculated using the point (median) posterior estimates and provided as supplementary information.

**Figure 6:**
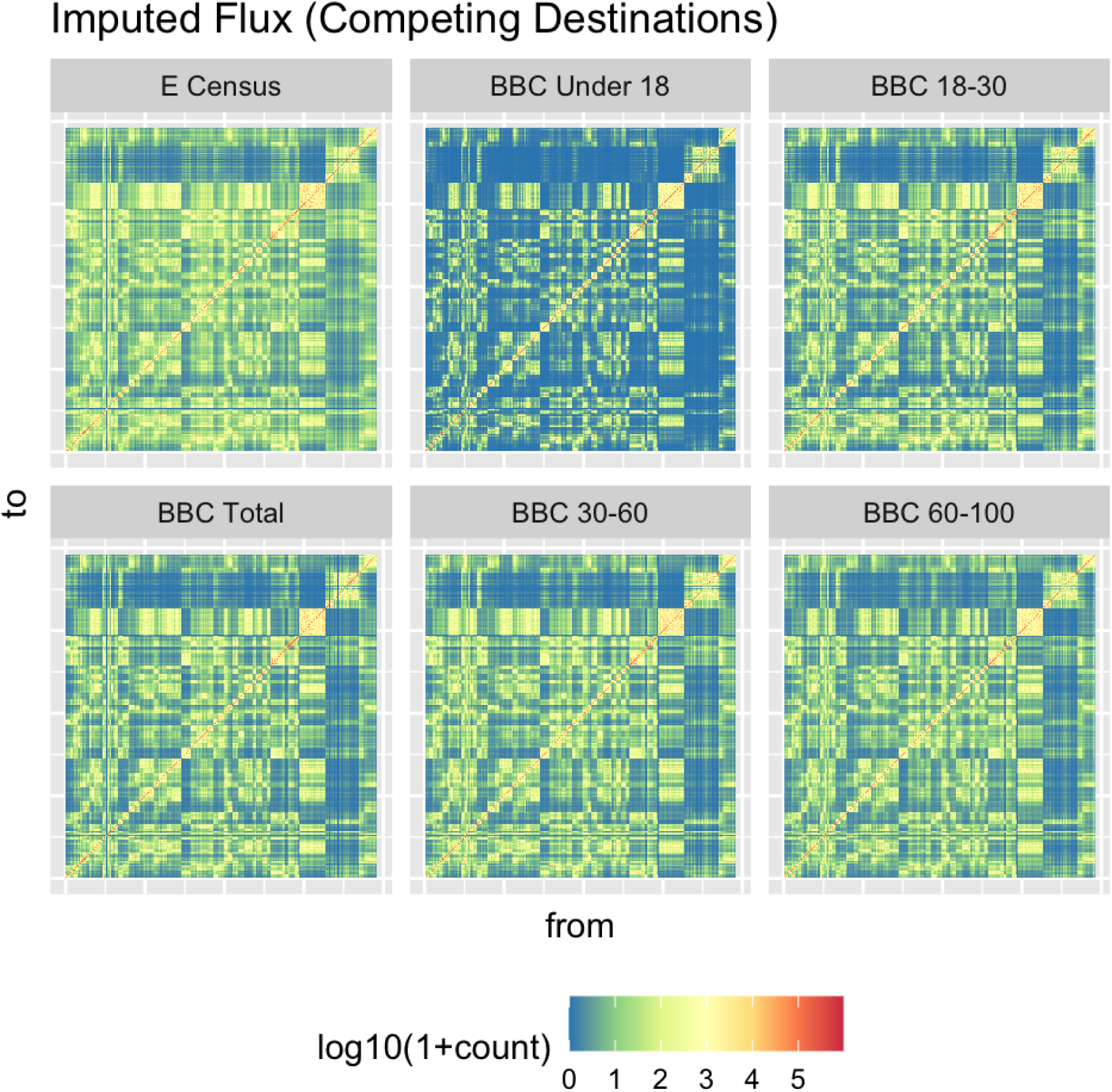
Imputed flux matrices for the Competing Destinations model. Imputed flux matrices from the Competing Destinations model (median parameter estimates) calculated for each age group of the BBC mobility data set (Total, Under 18, 18-30, 30-60 and 60-100) and compared to the imputed flux from the England census data (predicted to the UK demography).

### 2.7 Force of infection for a multi-group commuter model

To explore the extent to which variation in commuting rates and patterns within a population translates to epidemiological risk we derive a commuter approximation for the force of infection for a generic SIR (susceptible *S*, infected *I*, recovered *R*) model within a meta-population with multiple movement groups. We assume that the force of infection is well mixed within each patch. The effective local force of infection within a patch can be conceptually thought of as being constituted by two parts: the local force of infection due to resident infectives and the extrinsic force of infection generated by infected movers resident within the local population and susceptible movers to other spatial locations.

Elaborating an earlier result from [27], [7] demonstrated that the magnitude of these two components can be explicitly derived from the mechanistic movement rates. The per capita movement rate 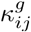 from patch *i* to patch *j* for members of group *g* can be calculated in terms of the inferred probability of movement 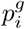 and conditional movement matrix 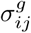 :

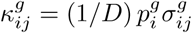

where *D* is the average trip duration.

The total movement rate from patch *i* to patch *j* will then be:

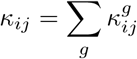

As long as the return rate 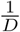 is small with respect to the generation time of the pathogen then the force of infection acting on susceptibles (*S*_*ig*_) in patch *i* and movement group *g* can be approximated as:

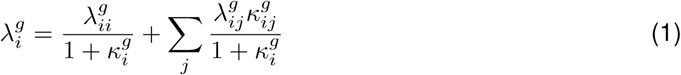

where 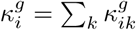.

We further assume that individuals differ only with respect to their mobility and have equal susceptibility and infectiousness and mix homogeneously with other individuals within the same patch. Under this assumption the force of infection only depends on the total number of infected individuals in each patch: *I*_*i*_ = Σ_*g*_ *I*_*ig*_. The first term corresponds to the contribution to the force of infection acting on susceptibles within group *g* resident in patch *i*:

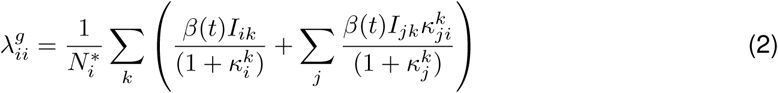

This has also two terms, the first corresponds to the contribution from local infectives (who are not commuting) from all four mobility groups. The second term corresponds to the contribution of susceptible individuals from patch *j* meeting infectious individuals (from all mobility groups *g*) while commuting. Similarly, the contribution of local susceptibles encountering infectious individuals (of all groups) while commuting to another patch is:

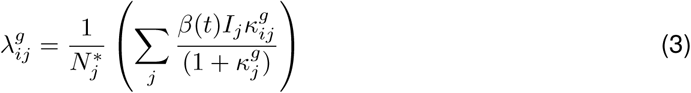

The effective population size within each patch is:

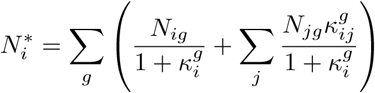

where the population size *N*_*i*_ = Σ_*g*_ *S*_*ig*_ + *I*_*ig*_ + *R*_*ig*_ and we neglect higher order terms of the movement matrix *O*(*σ*^2^) and higher.

### 2.8 Geographic risk of transmission

We use our multi-group commuter model to explore how the predicted geographic risk of transmission differs between an aggregate and age-stratified commuter model. Previous theoretical work has suggested that difference in mobility rates (or equivalently average trip duration *D*) are most important in the early stages of invasion when incidence is low [7]. As an illustration we consider an invasion seeded, as was the BBC pandemic, in Haslemere situated within the Waverley district in southern England. We consider the scenario where a novel pathogen has been introduced into a single location and identify the age-group that contributes the largest value to the net force of infection assuming a single infectious individual within each age group (Figure 7 A). We note first that although modulated by population density (7 C), distance from the seed location is the primary determinant of the net contribution to the force of infection in distant LADs ((7 B). While young people – in this case the 18-30 group – provide the largest net risk of transmission to another LAD – the relative contribution of older age-groups becomes more important for distant, low density areas (7 B) which also tend to have older populations (7 D). In supplemental information we explore a range of different seeder locations in comparable commuter districts across the UK (Falkirk, Belfast, Newport) and see the same basic pattern with one small variation. For low density districts proximal to the seed location the youngest Under 18 year old group, whose movements are closely constrained to their home location, are occasionally the dominant group - perhaps reflecting the size of school catchment areas.

**Figure 7:**
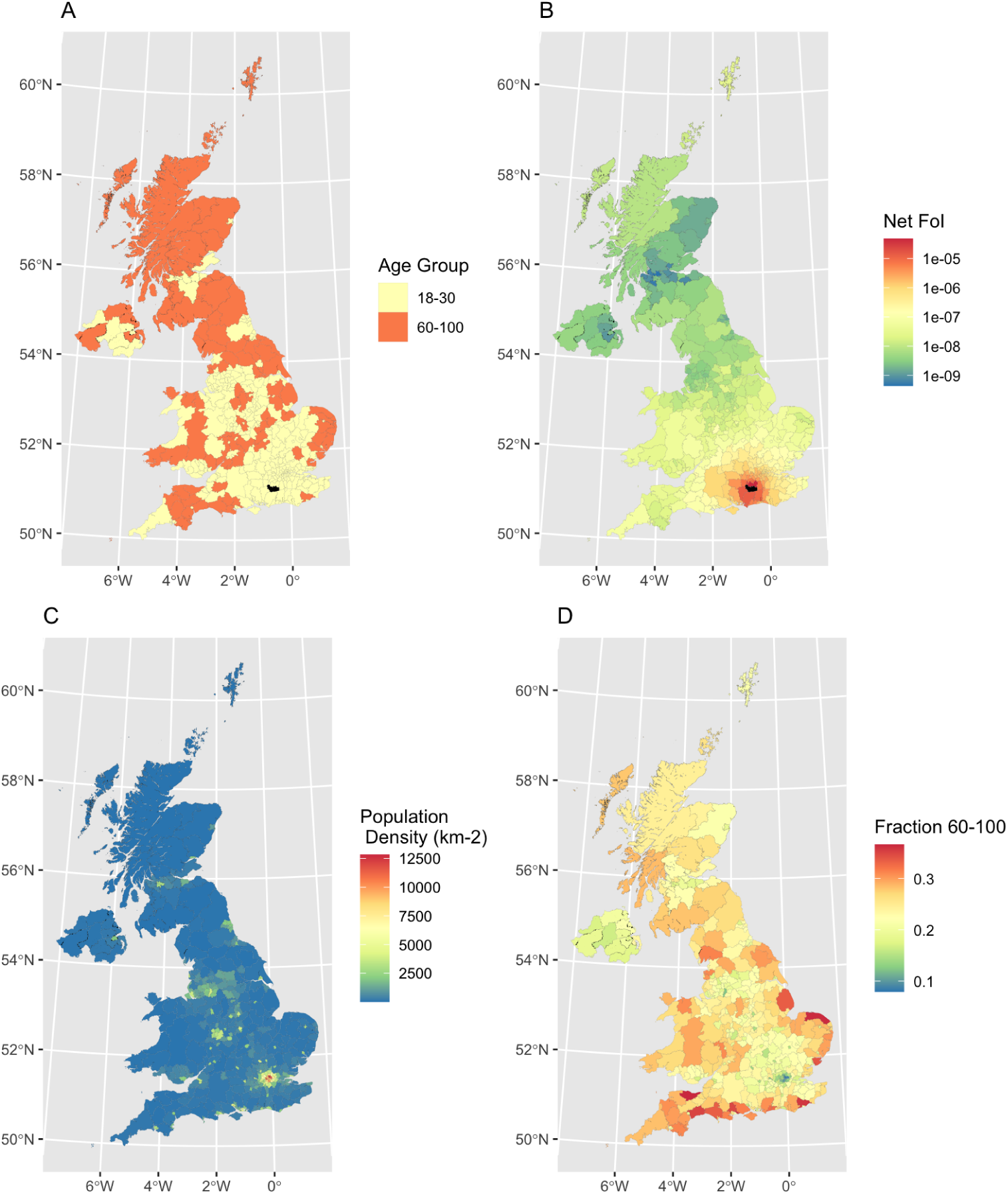
Dominant age group contributing to local force of infection. Set of choropleth maps exploring the predicted force-of-infection across the UK from a single infectious individual within each age-group located in Waverely (LAD containing Haslemere, filled black on maps **A,B**) compared with demographic correlates (population density and fraction of population aged 60-100). Panel **A** The age-group with the largest contribution to the local force of infection with each LAD. Panel **B** The net force of infection within each LAD (sum over contribution from all age groups). Panel **C** Local population density within each LAD (Total population size per *km*^2^). Panel **D** Fraction of local population in 60-100 year old age group. While the fall-off in the net force of infection is driven primarily by distance - the relative contribution of different age groups is shaped by population density and demography with individuals in the 60-100 age range playing the dominant role in transmission to low density areas with older populations, while the 18-30 year group is more important for cities which also have younger populations.

## 3 Discussion

In this paper we have introduced mobility data from the BBC Pandemic project to perform an analysis of how commuting patterns differ between groups of individuals with respect to employment status. We estimate and compare seven alternative human mobility models and find that the Competing Destinations model (with offset distance kernel, CDO) provides the best fit and predictive ability as assessed by leave-one-out (LOO) cross validation and posterior predictive checks of the common part of commuters (CPC) index. The competing destinations model extends the classical gravity formulation by adding a term that adjusts the flux between two locations according to the network of alternative locations within the meta-population. Although it lacks the elegant theoretical origins and mechanistic interpretation of the Radiation model – the additional flexibility makes it more appropriate for exploring the differences in mobility patterns between different groups.

However we note that, despite requiring two less parameters than the favoured CDO model, the Extended radiation model has similar absolute predictive performance in terms of the common part of commuters (CPC) index. The estimated value of *α* = 0.49 (0.48 *−* 0.5) for the full UK BBC Total data set is in line, but slightly higher than, the expected value of *α* = 0.43 independently estimated by McNeill et al. [36] based on a length scale for average trips of *l* = 19*km*.

Users in the BBC mobility data set demonstrate a higher rate of mobility across employment categories. There are important differences in the rates of mobility between urban and rural areas for users in different employment and age groups which could potentially be important for modelling the invasion of novel pandemic pathogens as the increased mobility and range of older individuals and those outside employment could enhance the rate of spread in the early stages of an outbreak.

For this paper we constructed origin-destination flows from users GPS trajectories taking the most frequent location as the inferred home (origin) and considered two alternative measures for the destination. The results we present in this paper use the second most frequent location (“next”) as the users destination. Repeating the analysis using the furthest extent and all recorded user locations (other than the origin) give the same qualitative results in terms of the relative performance of mobility models and differences between employment categories. We chose to focus on the “next” definition given it’s logical consistency with the question asked in the UK census. To validate this assumption by comparing the common part of commuters (CPC) between the predicted flux from the England & Wales census data to the flux predicted by the best fit models to the BBC Total data (Figure 8). Both definitions lead to estimated models with predicted flux that has a high (and indistinguishable) degree of similarity to the flux predicted from models estimated from Census data. For our purposes in this paper, to explore how human mobility between different employment groups in society varies from patterns inferred from census data, it is clearly the most appropriate choice. However, it is not necessarily the most appropriate choice for predicting disease transmission, hence we include both alternative definitions and estimated models within the the on-line data respository.

**Figure 8:**
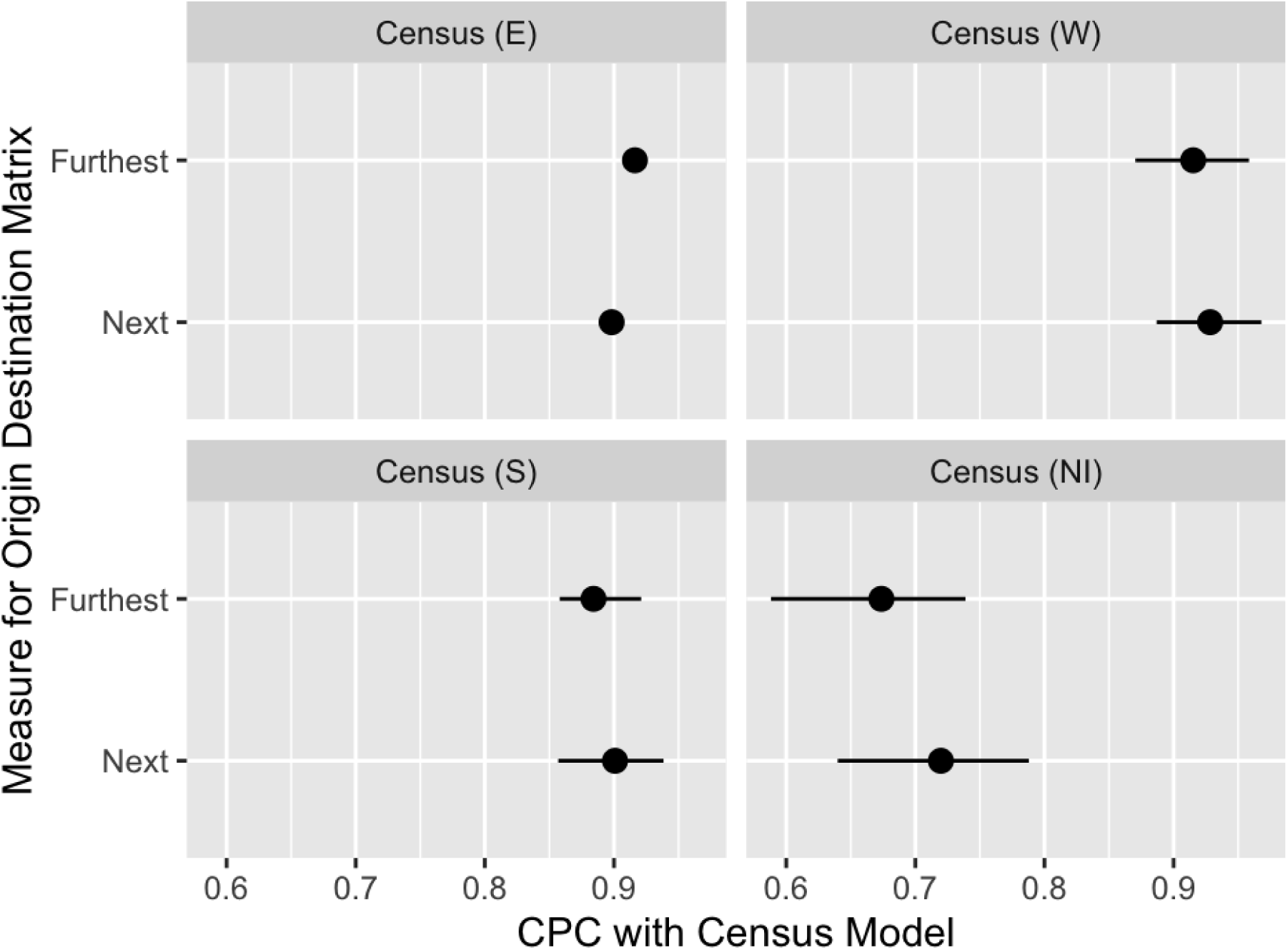
Comparison of posterior predicted flux from Census model to BBC mobility for different measures of destination. We use the common part of commuters (CPC) measure to assess which destination measure is most consistent with Census flux data. Flux was predicted for the whole of the UK using samples from the competing destinations model (CDO) estimated from the English census data and compared to the predicted flux for the same model estimated from the BBC Total data set constructed using the **next** most frequent and **furthest** extent definitions for destination location. The two measures are indistinguishable in terms of the difference in predictive accuracy compared to the equivalent model estimated from census data from the four member countries of the UK.

An unavoidable consequence of the crowd-sourced nature of our data is that users knew when their movements were being recorded and chose when to start tracking on the app which could potentially have changed their behaviour. We expect that this effect will be mitigated to an extent by the vast majority of users that signed up on (or immediately after) the day of broadcast. Although recruitment through the app was open over the course of a year, the largest group of users signed up following the broadcast of the documentary. There was also a smaller wave of recruitment following a social media campaign before filming was carried out in the town of Haslemere. This temporal pattern highlights both the effectiveness of the documentary and public engagement activities on recruitment, but also the potential for bias to be introduced into our sample of user trajectories. The close correspondence between the predicted flux and gravity parameters from the Total BBC data and subnational census workflows provides another layer of reassurance about the representativeness of the BBC data.

Census workflow data (and privately held mobile data sets) have the advantage of being dense enough that the raw data can be used directly in commuter models [5] without the need to estimate the human mobility models necessary to interpret the BBC mobility data. The consistency of the total BBC data at the aggregate level to census data, belies the differences in both the probability of movement (from home) and the choice of destination for movers not in full time employment. The unique meta-data collected along with GPS traces from the BBC Pandemic app has allowed us to quantify these differences for the first time. Under 18s and the over 60s are both less likely to move and have destinations closer to home than those in employment. We used a multi-group commuter model to illustrate how the predicted spatial risk of transmission varies according to seeder cases in different age groups. These differences may be small in aggregate but could be critically important in assessing the risk of spillover between regions in the early stages of a pandemic or in the period immediately following the easing of lockdown restrictions.

## Data Availability

Anonymised data and all code required to replicate analyses within the paper are available from a online repository linked below.

https://github.com/BBCPandemic/BBCMobility

## A Definition of Origin-Destination Matrices

User locations were first snapped to the nearest MSOA based on the generalised (20m resolution) shape file provided by the Open Geography portal from the Office of National Statistics (ONS). Locations outside of the boundary of any MSOA were either excluded or snapped to the MSOA with the greatest area of overlap within a 1km buffer centered around the user location. A home (origin) location was defined for each user as their modal MSOA. In defining the duration of time spent in a location we needed to account for missing location logs for some users who moved into an area with poor service, or switched off their phone, during the observation period. For such gaps we make the assumption the user remained in the last seen location until a new location was logged and use the duration of time within each location to calculate the modal location. In the event of a tie we chose the location with the least amount of time spent in the 12 hours between 7am and 6pm (inclusive). Users to which we could not assign a home location were removed from the data set for analysis.

For destination locations, two alternative definitions were considered - the furthest extent and the second most frequent recorded location after home (which we will refer to as the ‘next’ location for convenience). For the ‘next’ location we again ranked locations based on the total duration of time spent including the inferred location between gaps as described above. In the event of ties the location with the greater proportion of time between 7am and 6pm was chosen. Once again, users to whom we could not assign a unique destination location were removed from the data set. In total there were 4,450 users we could not assign a unique origin and destination location according these definitions leaving a total of 43,291 users within the final BBC mobility data set.

For users with all location records in the same MSOA, their home and destination locations are both set to this unique value. As the LAD origin and destinations are mapped from these MSOA locations, at the LAD level, users can therefore have the same inferred origin and destination locations (even though they have moved between different MSOAs over the course of the reporting period).

## B Supplemental Figures

**Figure 9:**
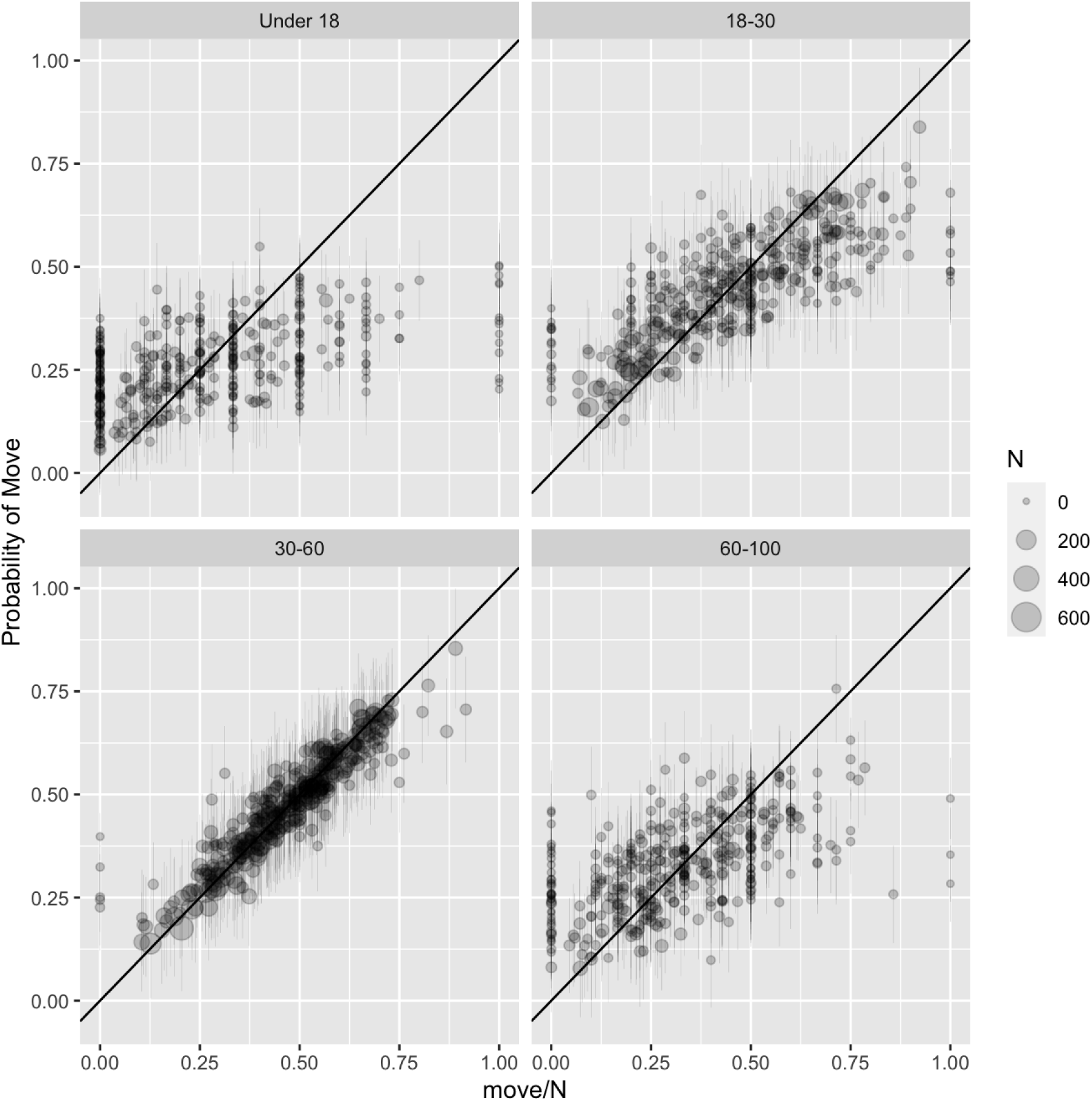
Predicted probability of movement from (age) random effects model. Predicted per capita probability of moving (i.e. having a different origin and destination location) from estimated random effects model plotted against observed proportion from BBC mobility data set (move/N). Error bars indicate 95% bootstrapped prediction intervals, size the number of observations (N) and the 1:1 line is added for reference.

**Figure 10:**
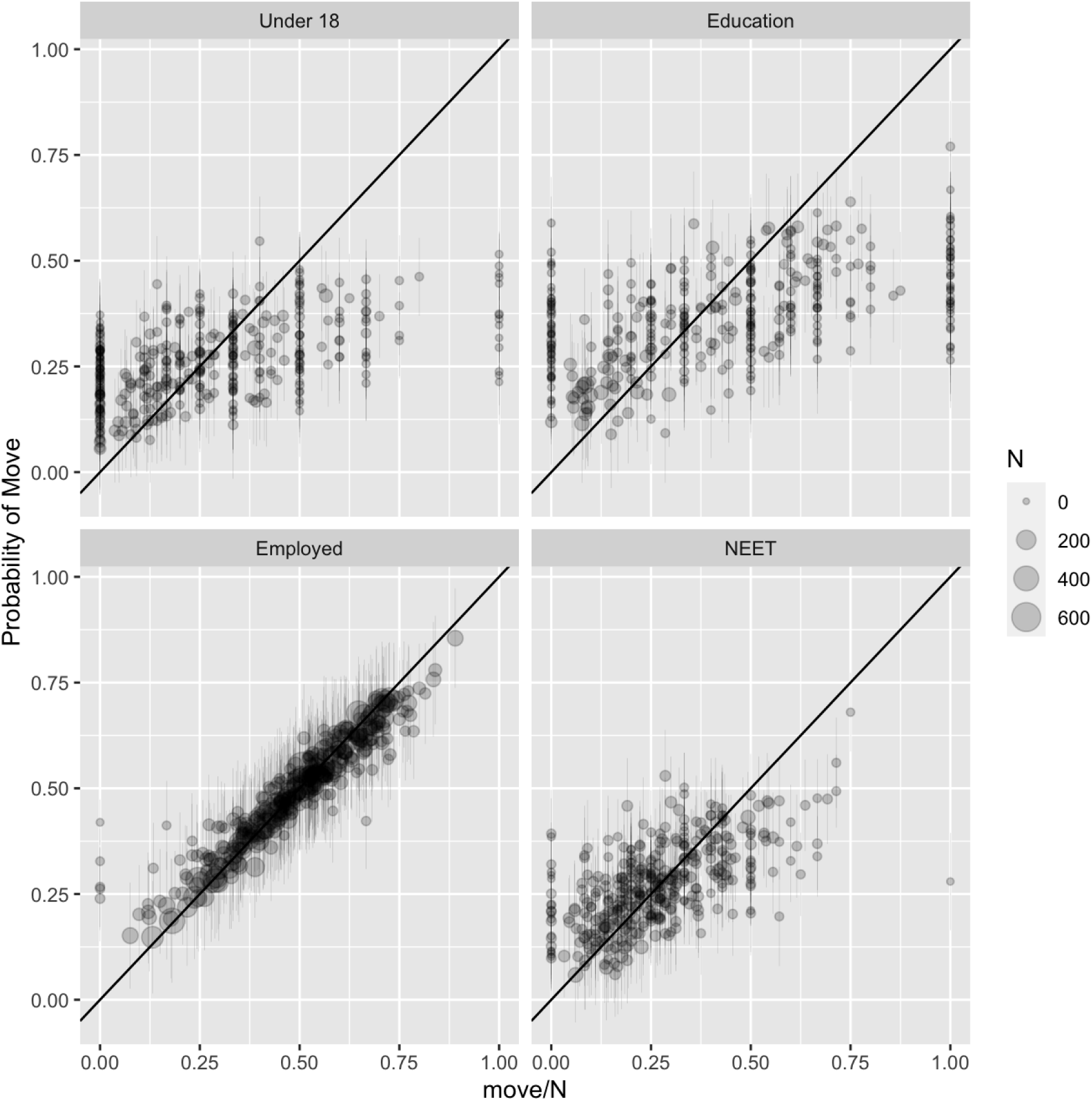
Predicted probability of movement from (employment) random effects model. Predicted per capita probability of moving (i.e. having a different origin and destination location) from estimated random effects model plotted against observed proportion from BBC mobility data set (move/N). Error bars indicate 95% bootstrapped prediction intervals, size the number of observations (N) and the 1:1 line is added for reference.

**Figure 11:**
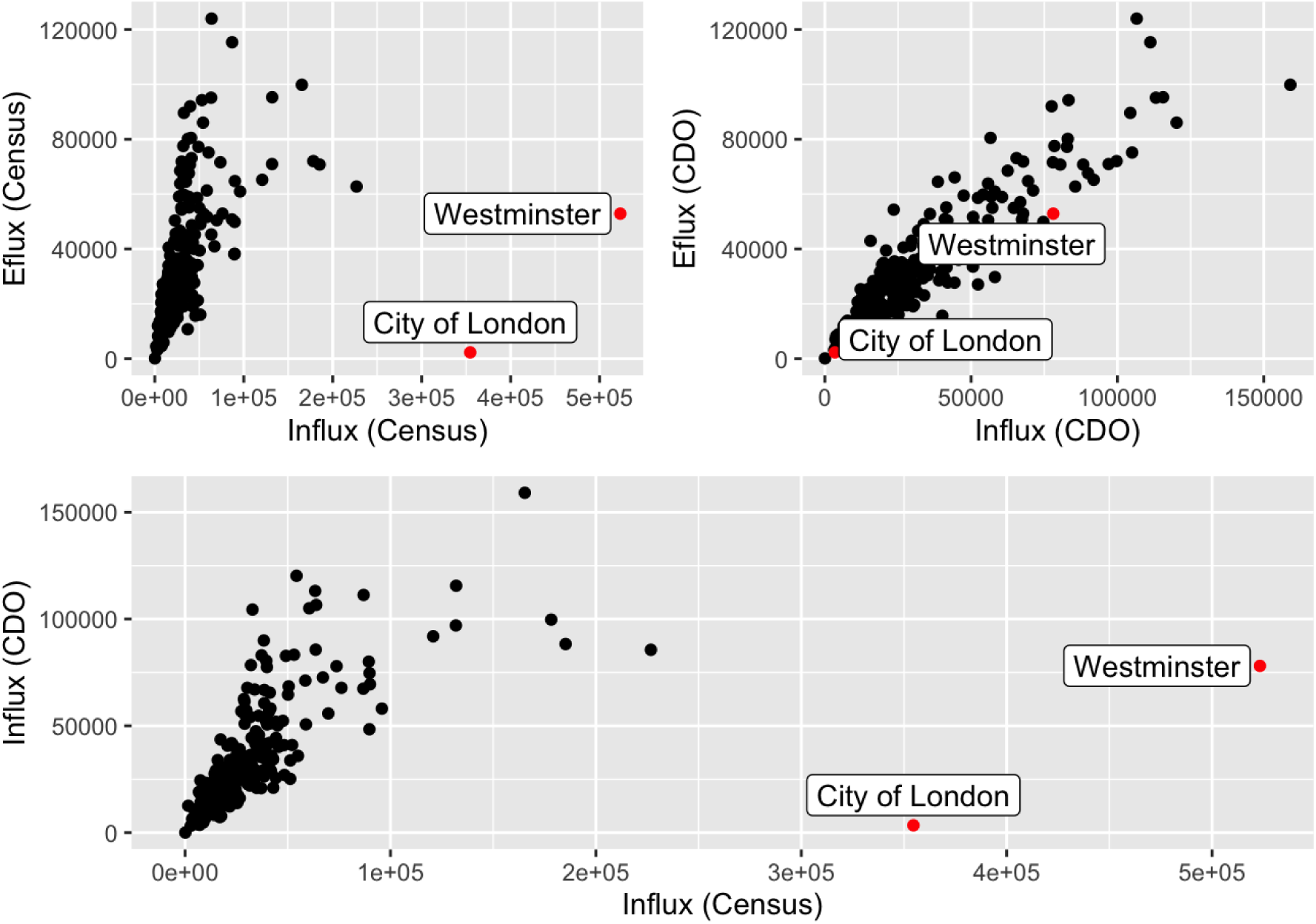
Influx and efflux of commuters in English Census Workflow Data. The City of London and Westminster (and to a lesser extent other boroughs of London) attract anomalously large numbers of commuters for their size. We visualise this by plotting the numbers of commuters in (influx) against leaving (efflux). For the majority of LADs this relationship is symmetric (top left panel), however the City of London and Westminster have orders of magnitude higher commuters in than residents who commute out. This flux is not captured by gravity type models as illustrated by the predicted flux from the CDO model (point prediction from median parameters, top right panel) and comparison of the empirical influx of commuters to the CDO prediction (bottom panel).

**Figure 12:**
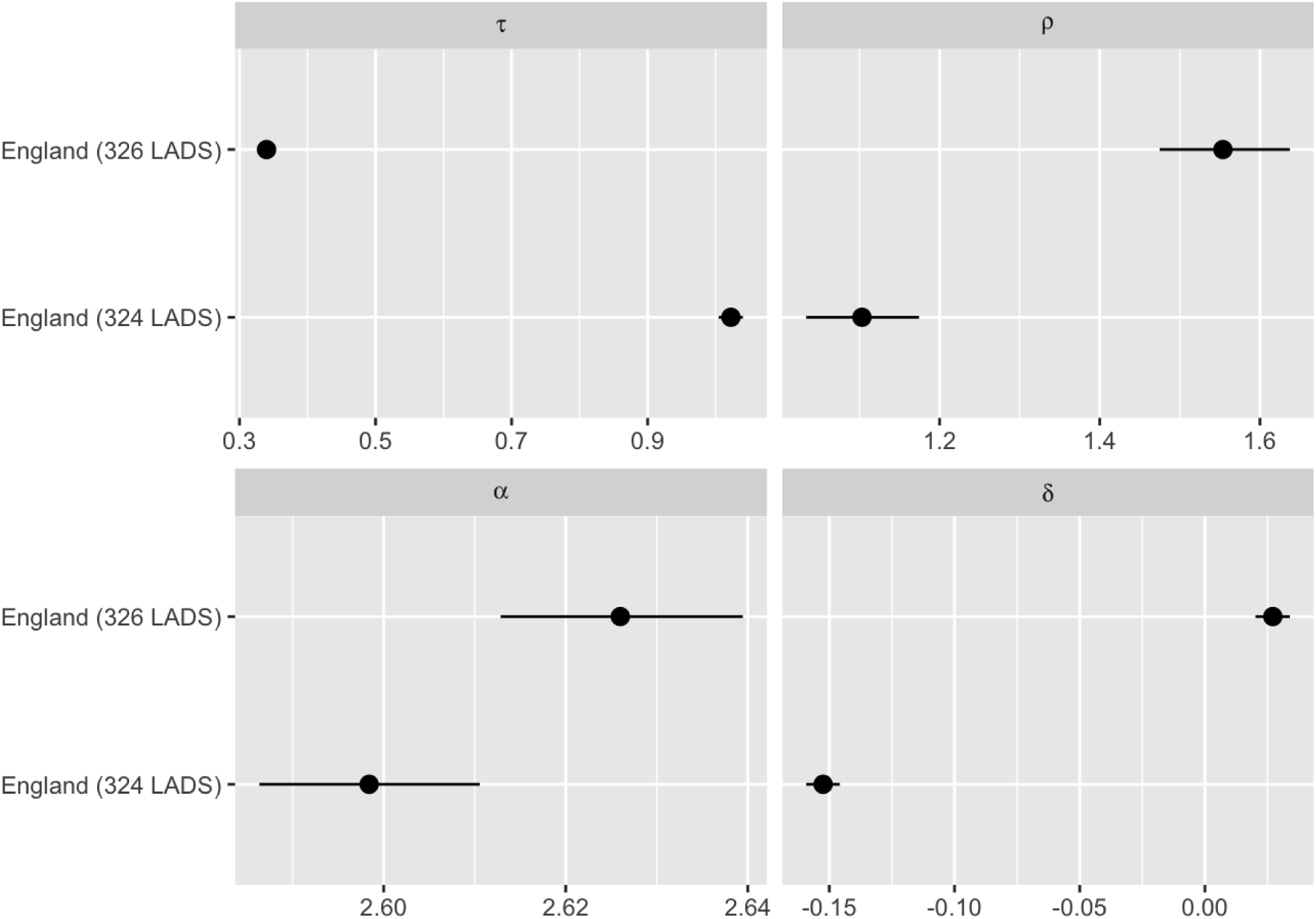
Systematic bias to parameter estimates of CDO model. Comparison of gravity model (CDO) parameter estimates (median and 95% credible intervals) from English census workflow data from the full 326 LADs and a reduced data set (324 LADS) removing the City of London and Westminster.

**Figure 13:**
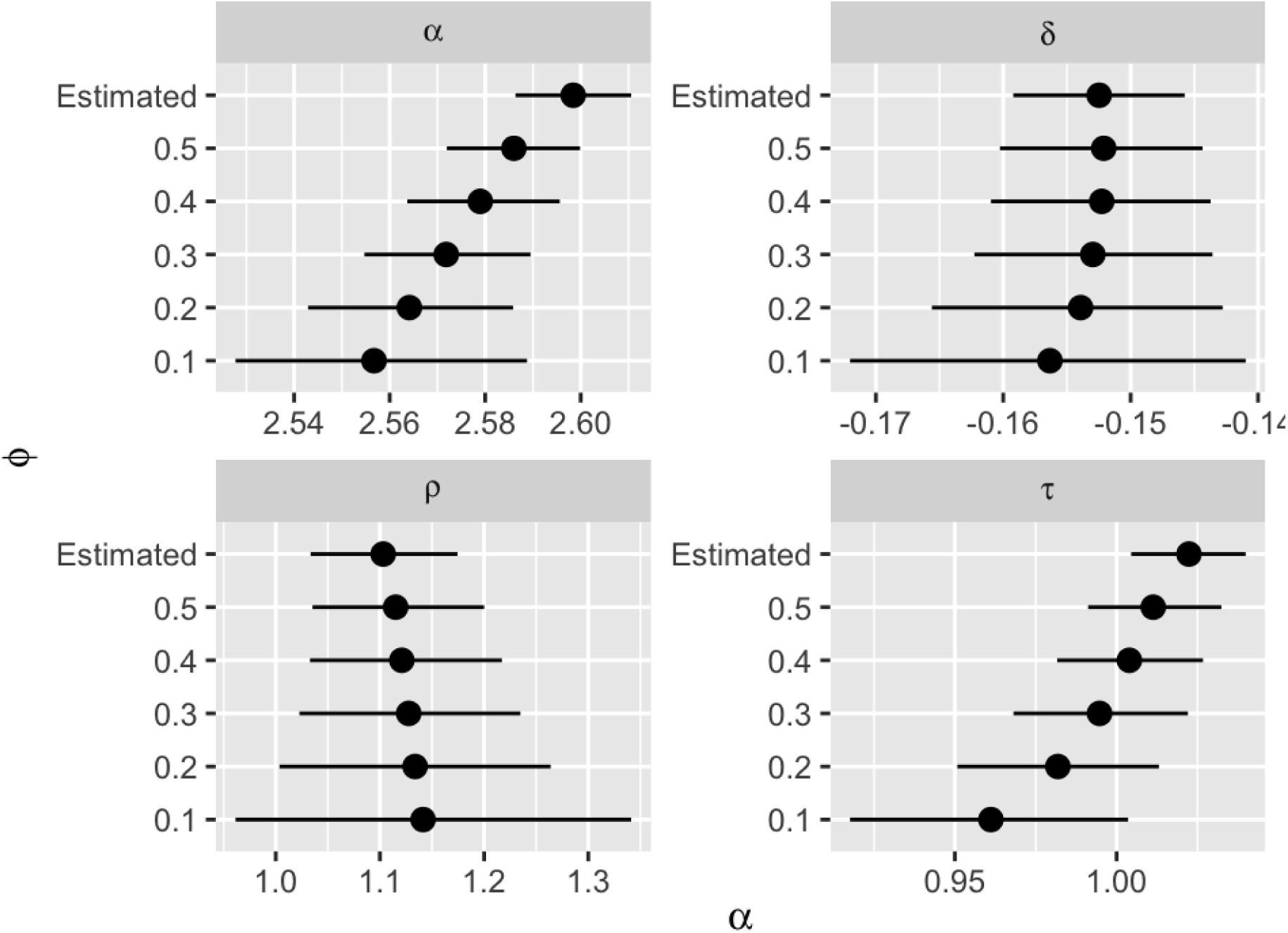
Change in posterior estimates of CDO model with fixed shape parameter (*ϕ*) Comparison of estimated posterior distributions for the CDO gravity scaling parameters estimated from the English census workflow data (324 LADs) for different (fixed) values of the shape parameter *ϕ* compared to the estimated value.

## C Figure supplements to Figure 7

**Figure 14:**
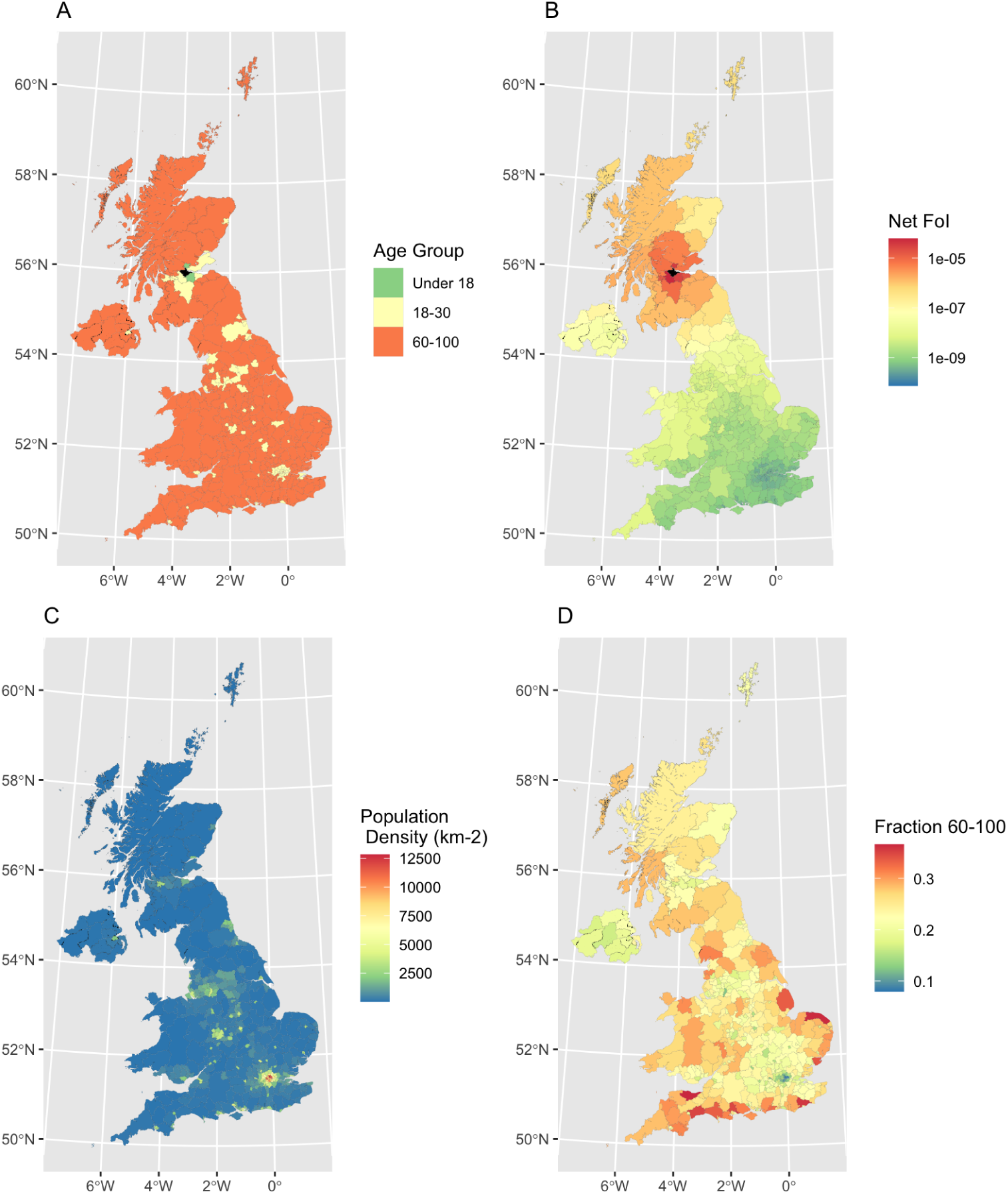
Dominant age group contributing to local force of infection. Set of choropleth maps exploring the predicted force-of-infection across the UK from a single infectious individual within each age-group located in the Scottish commuter belt town of Falkirk (filled black on maps **A,B**) compared with demographic correlates (population density and fraction of population aged 60-100). Panel **A** The age-group with the largest contribution to the local force of infection with each LAD. Panel **B** The net force of infection within each LAD (sum over contribution from all age groups). Panel **C** Local population density within each LAD (Total population size per *km*^2^). Panel **D** Fraction of local population in 60-100 year old age group. While the fall-off in the net force of infection is driven primarily by distance - the relative contribution of different age groups is shaped by population density and demography. The 18-30 year group dominates for cities and high density LADs, which also tend to have younger populations, while individuals in the 60-100 age group play the dominant role in transmission to low density areas. Under 18s play a more important role for low-density areas when they are proximal to the seed location – in this case Clackmannanshire and West Lothian.

**Figure 15:**
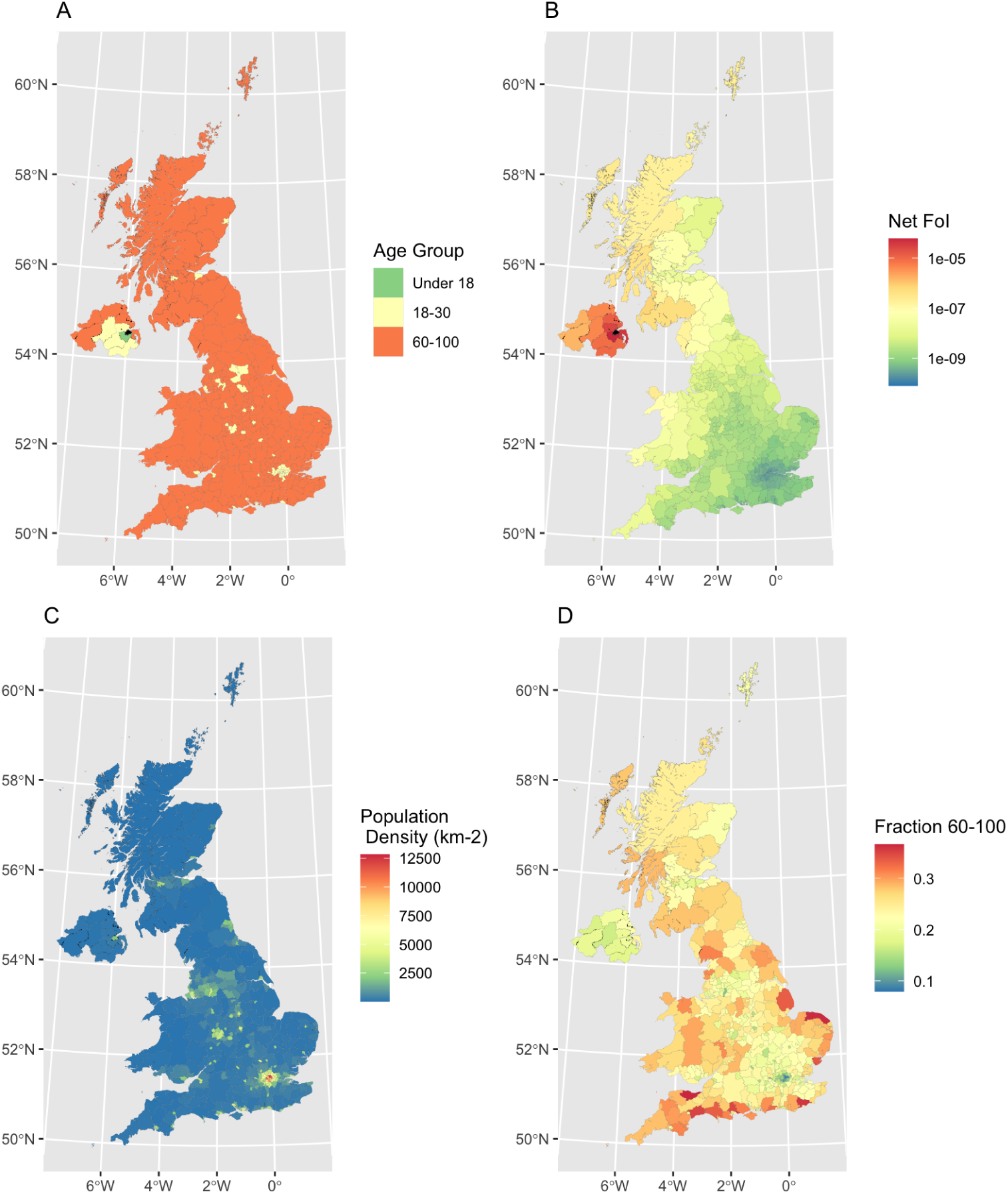
Dominant age group contributing to local force of infection. Set of choropleth maps exploring the predicted force-of-infection across the UK from a single infectious individual within each age-group located in Belfast, Northern Ireland (filled black on maps **A,B**) compared with demographic correlates (population density and fraction of population aged 60-100). Panel **A** The age-group with the largest contribution to the local force of infection with each LAD. Panel **B** The net force of infection within each LAD (sum over contribution from all age groups). Panel **C** Local population density within each LAD (Total population size per *km*^2^). Panel **D** Fraction of local population in 60-100 year old age group. While the fall-off in the net force of infection is driven primarily by distance - the relative contribution of different age groups is shaped by population density and demography. The 18-30 year group dominates for cities and high density LADs, which also tend to have younger populations, while individuals in the 60-100 age group play the dominant role in transmission to low density areas. Under 18s play a more important role for low-density areas when they are proximal to the seed location – in this case Lisburn and Castlereagh.

**Figure 16:**
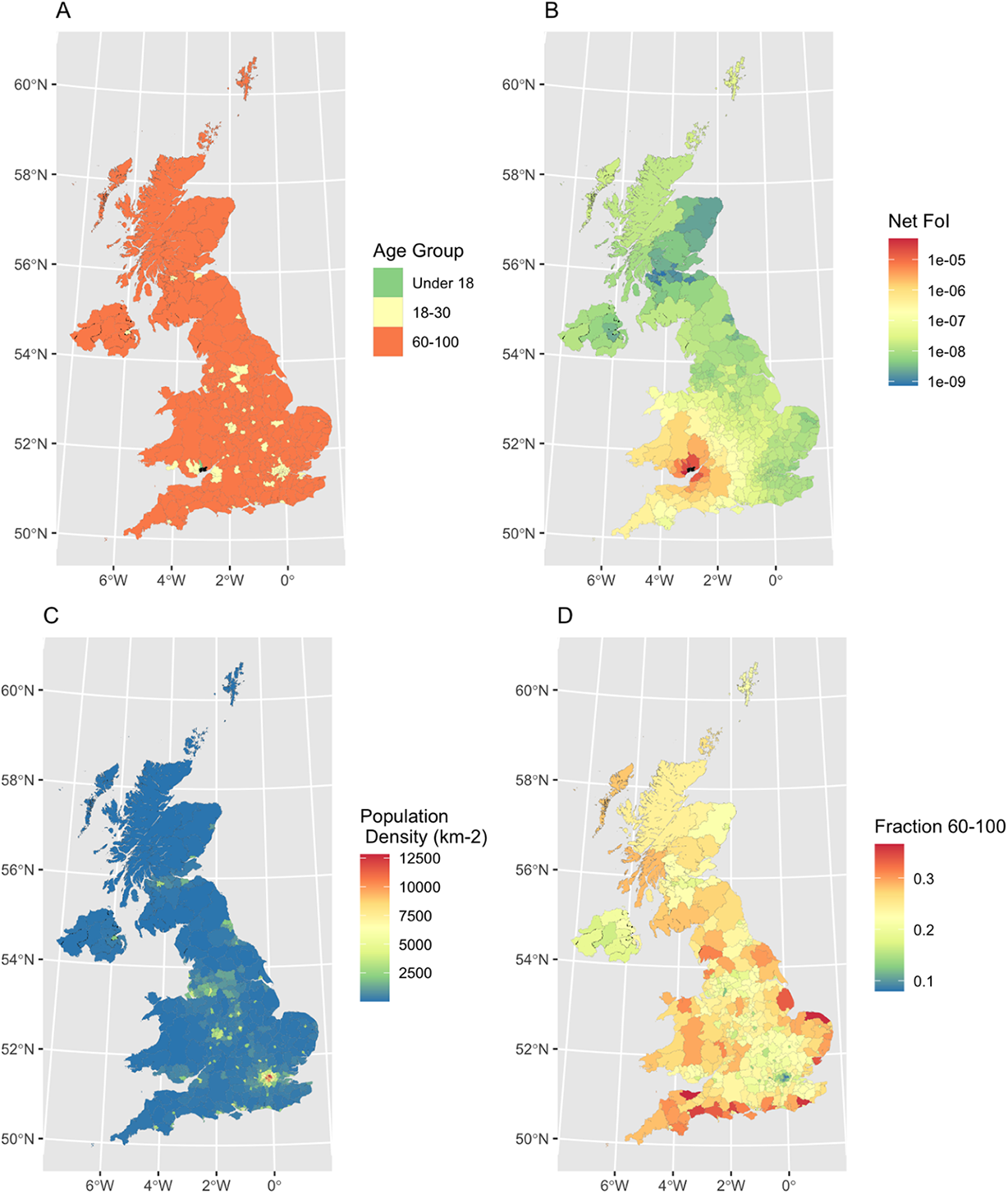
Dominant age group contributing to local force of infection. Set of choropleth maps exploring the predicted force-of-infection across the UK from a single infectious individual within each age-group located in Newport, Wales (filled black on maps **A,B**) compared with demographic correlates (population density and fraction of population aged 60-100). Panel **A** The age-group with the largest contribution to the local force of infection with each LAD. Panel **B** The net force of infection within each LAD (sum over contribution from all age groups). Panel **C** Local population density within each LAD (Total population size per *km*^2^). Panel **D** Fraction of local population in 60-100 year old age group. While the fall-off in the net force of infection is driven primarily by distance - the relative contribution of different age groups is shaped by population density and demography. The 18-30 year group dominates for cities and high density LADs, which also tend to have younger populations, while individuals in the 60-100 age group play the dominant role in transmission to low density areas. Under 18s play a more important role for low-density areas when they are proximal to the seed location – in this case Rhondda Cynon Taf.

**Figure 17:**
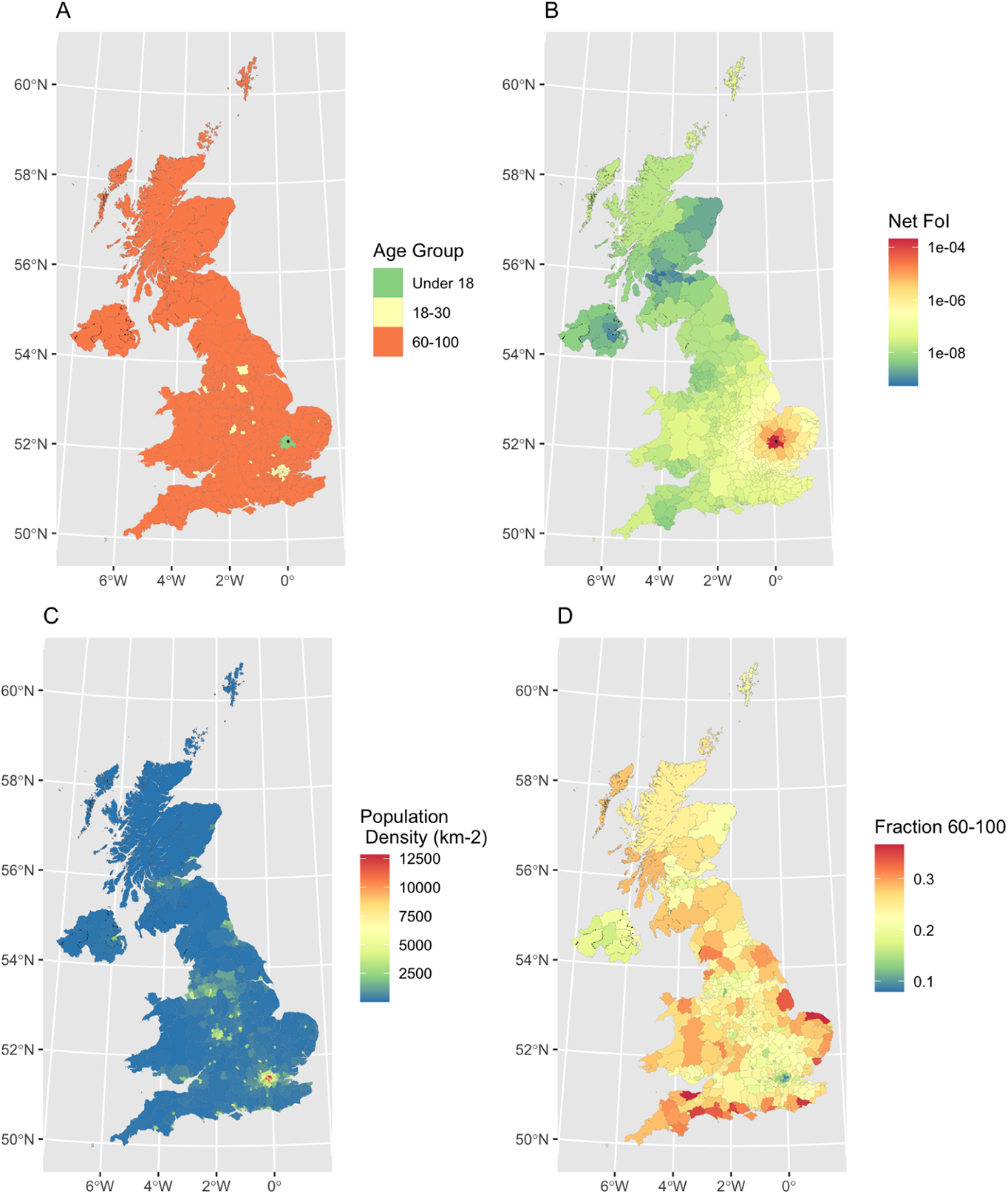
Dominant age group contributing to local force of infection. Set of choropleth maps exploring the predicted force-of-infection across the UK from a single infectious individual within each age-group located in Cambridge (filled black on maps **A,B**) compared with demographic correlates (population density and fraction of population aged 60-100). Panel **A** The age-group with the largest contribution to the local force of infection with each LAD. Panel **B** The net force of infection within each LAD (sum over contribution from all age groups). Panel **C** Local population density within each LAD (Total population size per *km*^2^). Panel **D** Fraction of local population in 60-100 year old age group. While the fall-off in the net force of infection is driven primarily by distance - the relative contribution of different age groups is shaped by population density and demography. The 18-30 year group dominates for cities and high density LADs, which also tend to have younger populations, while individuals in the 60-100 age group play the dominant role in transmission to low density areas. Under 18s play a more important role for low-density areas when they are proximal to the seed location – in this case the surrounding rural district of South Cambridgeshire.

**Figure 18:**
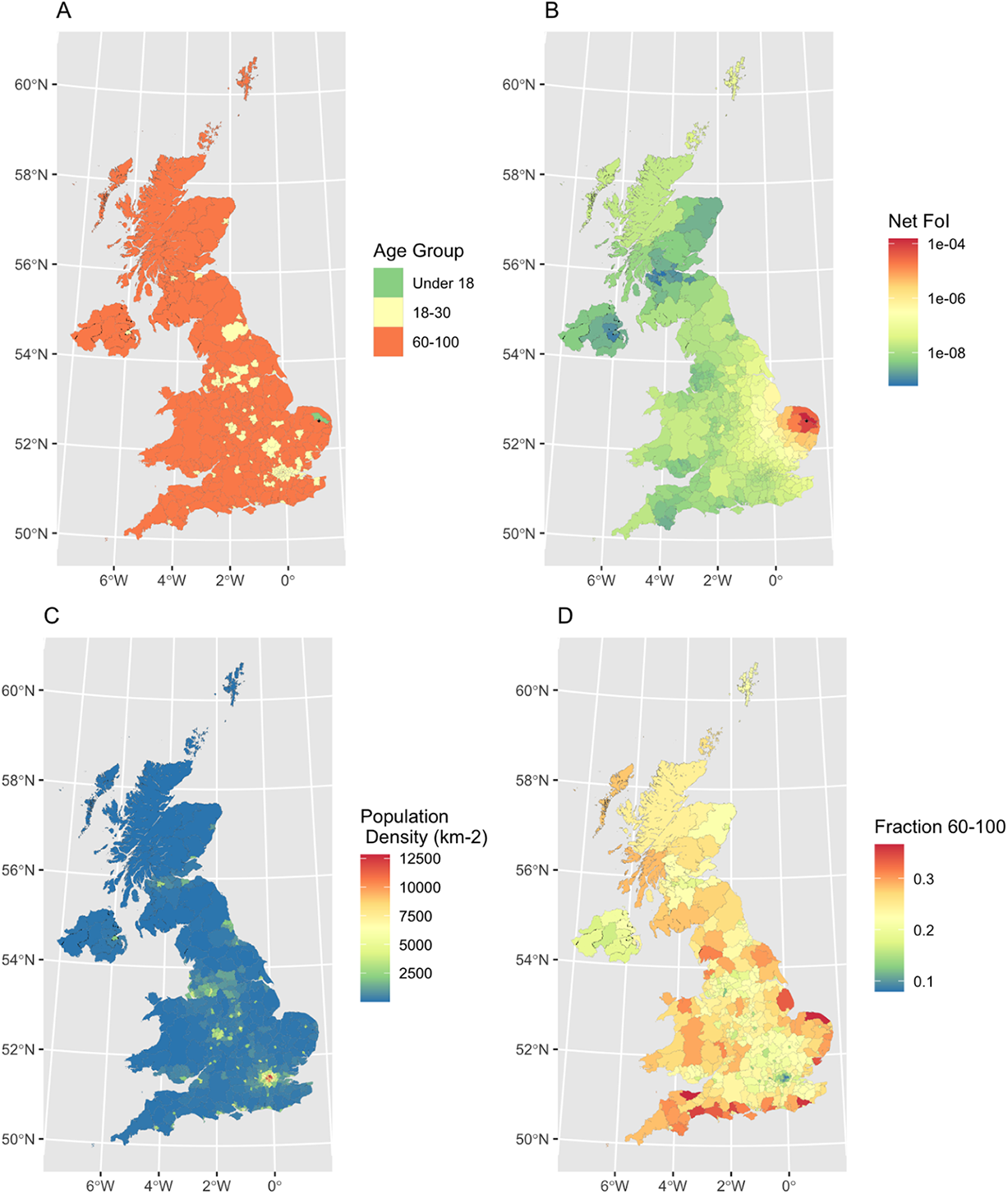
Dominant age group contributing to local force of infection. Set of choropleth maps exploring the predicted force-of-infection across the UK from a single infectious individual within each age-group located in Norwich (filled black on maps **A,B**) compared with demographic correlates (population density and fraction of population aged 60-100). Panel **A** The age-group with the largest contribution to the local force of infection with each LAD. Panel **B** The net force of infection within each LAD (sum over contribution from all age groups). Panel **C** Local population density within each LAD (Total population size per *km*^2^). Panel **D** Fraction of local population in 60-100 year old age group. While the fall-off in the net force of infection is driven primarily by distance - the relative contribution of different age groups is shaped by population density and demography. The 18-30 year group dominates for cities and high density LADs, which also tend to have younger populations, while individuals in the 60-100 age group play the dominant role in transmission to low density areas. Under 18s play a more important role for low-density areas when they are proximal to the seed location – in this case the adjacent district of Broadland.

## D Posterior estimates

**Table 8:**
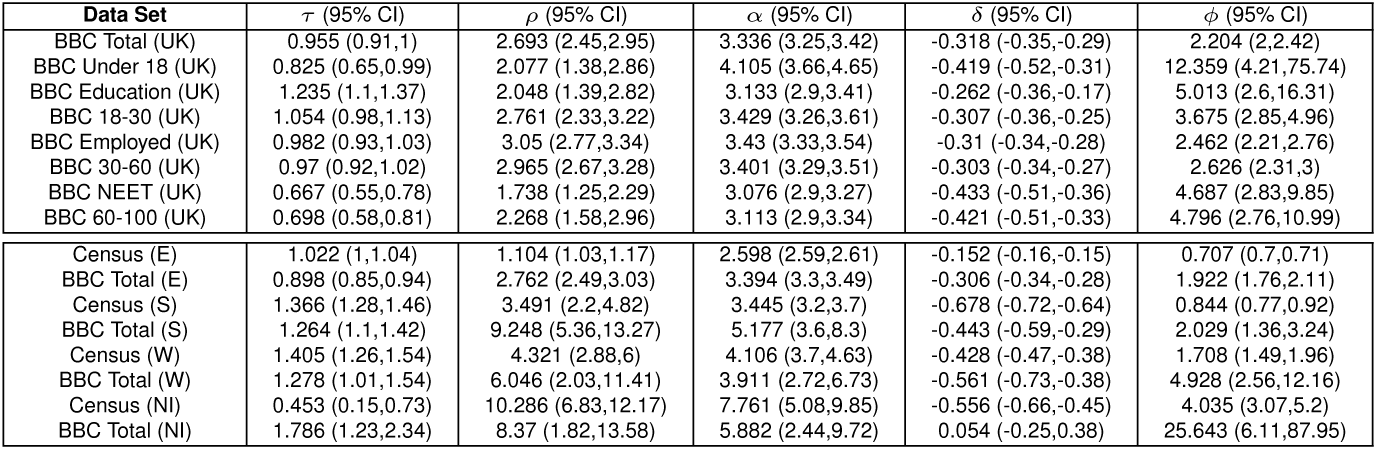
Posterior estimates for Competing Destinations (CDO) model.

**Table 9:** Posterior estimates for Competing Destinations (CDP) model.

| Data Set | $\tau$ (95% CI) | $\rho$ (95% CI) | $\delta$ (95% CI) | $\phi$ (95% CI) |
| --- | --- | --- | --- | --- |
| BBC Total (UK) | 0.919 (0.87,0.96) | 2.547 (2.52,2.57) | -0.261 (-0.28,-0.24) | 1.757 (1.61,1.92) |
| BBC Under 18 (UK) | 0.808 (0.64,0.98) | 3.037 (2.9,3.18) | -0.369 (-0.47,-0.27) | 7.157 (3,39.9) |
| BBC Education (UK) | 1.232 (1.1,1.37) | 2.512 (2.43,2.6) | -0.252 (-0.34,-0.17) | 3.829 (2.2,9.03) |
| BBC 18-30 (UK) | 1.032 (0.96,1.11) | 2.512 (2.46,2.56) | -0.259 (-0.31,-0.21) | 2.724 (2.18,3.48) |
| BBC Employed (UK) | 0.943 (0.9,0.99) | 2.525 (2.5,2.55) | -0.247 (-0.27,-0.22) | 1.889 (1.72,2.09) |
| BBC 30-60 (UK) | 0.936 (0.88,0.98) | 2.53 (2.5,2.56) | -0.243 (-0.27,-0.22) | 2.003 (1.79,2.26) |
| BBC NEET (UK) | 0.662 (0.55,0.78) | 2.566 (2.49,2.64) | -0.397 (-0.47,-0.33) | 3.595 (2.35,6.28) |
| BBC 60-100 (UK) | 0.695 (0.58,0.81) | 2.5 (2.43,2.57) | -0.381 (-0.46,-0.31) | 3.54 (2.24,6.73) |
| Census (E) | 1.042 (1.02,1.06) | 2.409 (2.4,2.41) | -0.162 (-0.17,-0.16) | 0.695 (0.69,0.7) |
| BBC Total (E) | 0.856 (0.81,0.9) | 2.534 (2.51,2.56) | -0.253 (-0.28,-0.23) | 1.571 (1.45,1.71) |
| Census (S) | 1.283 (1.19,1.37) | 2.829 (2.75,2.91) | -0.627 (-0.67,-0.59) | 0.813 (0.74,0.89) |
| BBC Total (S) | 1.22 (1.06,1.38) | 2.412 (2.22,2.62) | -0.371 (-0.51,-0.23) | 1.599 (1.11,2.41) |
| Census (W) | 1.289 (1.15,1.43) | 3.02 (2.95,3.09) | -0.435 (-0.47,-0.39) | 1.545 (1.35,1.77) |
| BBC Total (W) | 1.274 (0.99,1.55) | 2.395 (2.13,2.66) | -0.559 (-0.71,-0.4) | 4.287 (2.33,9.37) |
| Census (NI) | 0.356 (0.07,0.66) | 2.617 (2.42,2.79) | -0.536 (-0.65,-0.41) | 2.52 (1.94,3.2) |
| BBC Total (NI) | 1.72 (1.1,2.27) | 2.143 (1.67,2.64) | 0.089 (-0.2,0.45) | 21.31 (5.34,87.96) |

**Table 10:** Posterior estimates for Competing Destinations (CDE) model.

| Data Set | $\tau$ (95% CI) | $\rho$ (95% CI) | $\delta$ (95% CI) | $\phi$ (95% CI) |
| --- | --- | --- | --- | --- |
| BBC Total (UK) | 1.008 (0.95,1.06) | 23.156 (22.72,23.61) | -0.373 (-0.42,-0.33) | 0.784 (0.73,0.84) |
| BBC Under 18 (UK) | 0.828 (0.66,1) | 11.796 (11.06,12.62) | -0.509 (-0.64,-0.38) | 7.453 (2.84,58) |
| BBC Education (UK) | 1.16 (1.02,1.29) | 20.013 (18.89,21.26) | -0.275 (-0.4,-0.14) | 1.438 (0.99,2.2) |
| BBC 18-30 (UK) | 1.049 (0.97,1.13) | 18.962 (18.32,19.61) | -0.341 (-0.41,-0.27) | 1.416 (1.2,1.7) |
| BBC Employed (UK) | 1.013 (0.96,1.07) | 22.146 (21.71,22.57) | -0.368 (-0.42,-0.32) | 0.952 (0.88,1.03) |
| BBC 30-60 (UK) | 0.969 (0.91,1.02) | 22.085 (21.61,22.58) | -0.357 (-0.41,-0.31) | 0.995 (0.91,1.09) |
| BBC NEET (UK) | 0.635 (0.53,0.74) | 20.69 (19.79,21.68) | -0.493 (-0.6,-0.39) | 1.448 (1.07,2.06) |
| BBC 60-100 (UK) | 0.661 (0.55,0.78) | 20.978 (20.04,22.04) | -0.492 (-0.6,-0.38) | 1.91 (1.34,3.07) |
| Census (E) | 1.55 (1.53,1.57) | 54.482 (54.25,54.72) | -0.163 (-0.18,-0.14) | 0.386 (0.38,0.39) |
| BBC Total (E) | 0.954 (0.9,1.01) | 20.91 (20.5,21.31) | -0.35 (-0.4,-0.3) | 0.817 (0.76,0.88) |
| Census (S) | 1.611 (1.51,1.7) | 54.945 (52.19,58.01) | -0.867 (-0.92,-0.81) | 0.468 (0.43,0.51) |
| BBC Total (S) | 1.282 (1.12,1.44) | 21.131 (19.04,23.6) | -0.503 (-0.68,-0.32) | 1.809 (1.23,2.78) |
| Census (W) | 1.533 (1.35,1.69) | 24.186 (23.56,24.88) | -0.411 (-0.49,-0.33) | 1.178 (1.04,1.34) |
| BBC Total (W) | 1.285 (1.01,1.57) | 20.193 (17.5,23.74) | -0.559 (-0.77,-0.33) | 3.236 (1.79,6.89) |
| Census (NI) | 0.463 (0.13,0.78) | 18.159 (17.21,19.22) | -0.57 (-0.7,-0.44) | 3.752 (2.86,4.78) |
| BBC Total (NI) | 1.879 (1.32,2.44) | 14.7 (11.83,19.05) | 0.055 (-0.26,0.39) | 23.936 (4.81,90.56) |

**Table 11:** Posterior estimates for Extended Radiation (ERad) model.

| Data Set | $\alpha$ (95% CI) | $\phi$ (95% CI) |
| --- | --- | --- |
| BBC Total (UK) | 0.484 (0.47,0.5) | 1.408 (1.3,1.53) |
| BBC Under 18 (UK) | 0.634 (0.57,0.7) | 4.264 (2.12,18.15) |
| BBC Education (UK) | 0.481 (0.43,0.53) | 2.754 (1.72,5.33) |
| BBC 18-30 (UK) | 0.483 (0.46,0.51) | 2.041 (1.69,2.49) |
| BBC Employed (UK) | 0.471 (0.46,0.49) | 1.501 (1.37,1.64) |
| BBC 30-60 (UK) | 0.467 (0.45,0.48) | 1.606 (1.45,1.78) |
| BBC NEET (UK) | 0.45 (0.41,0.49) | 2.574 (1.77,4.2) |
| BBC 60-100 (UK) | 0.404 (0.37,0.44) | 2.265 (1.54,3.57) |
| Census (E) | 0.561 (0.56,0.56) | 0.918 (0.91,0.93) |
| BBC Total (E) | 0.467 (0.45,0.48) | 1.366 (1.26,1.48) |
| Census (S) | 0.476 (0.43,0.53) | 0.534 (0.49,0.58) |
| BBC Total (S) | 0.45 (0.34,0.57) | 1.016 (0.75,1.39) |
| Census (W) | 0.79 (0.71,0.87) | 0.533 (0.48,0.59) |
| BBC Total (W) | 0.553 (0.39,0.72) | 1.256 (0.85,1.92) |
| Census (NI) | 0.429 (0.26,0.59) | 1.399 (1.08,1.76) |
| BBC Total (NI) | 0.722 (0.45,0.95) | 12.453 (3.49,75.38) |

**Table 12:** Posterior estimates for Intervening Opportunities (IO) model.

| Data Set | $\log_{10}(\gamma)$ (95% CI) | $\phi$ (95% CI) |
| --- | --- | --- |
| BBC Total (UK) | -6.638 (-6.65,-6.63) | 0.377 (0.36,0.4) |
| BBC Under 18 (UK) | -6.243 (-6.28,-6.21) | 1.215 (0.79,2.06) |
| BBC Education (UK) | -6.538 (-6.57,-6.51) | 0.55 (0.42,0.72) |
| BBC 18-30 (UK) | -6.522 (-6.54,-6.5) | 0.66 (0.58,0.76) |
| BBC Employed (UK) | -6.617 (-6.63,-6.61) | 0.437 (0.41,0.47) |
| BBC 30-60 (UK) | -6.611 (-6.62,-6.6) | 0.439 (0.41,0.47) |
| BBC NEET (UK) | -6.549 (-6.57,-6.52) | 0.552 (0.44,0.7) |
| BBC 60-100 (UK) | -6.573 (-6.6,-6.55) | 0.621 (0.49,0.79) |
| Census (E) | -6.885 (-6.89,-6.88) | 0.424 (0.42,0.43) |
| BBC Total (E) | -6.611 (-6.62,-6.6) | 0.411 (0.39,0.43) |
| Census (S) | -6.019 (-6.04,-6) | 0.562 (0.52,0.61) |
| BBC Total (S) | -5.951 (-6,-5.91) | 1.065 (0.78,1.49) |
| Census (W) | -5.699 (-5.72,-5.68) | 0.588 (0.52,0.66) |
| BBC Total (W) | -5.725 (-5.78,-5.67) | 1.478 (0.97,2.32) |
| Census (NI) | -5.662 (-5.71,-5.62) | 1.77 (1.37,2.25) |
| BBC Total (NI) | -5.517 (-5.61,-5.44) | 19.444 (4.81,80.31) |

**Table 13:** Posterior estimates for Stoufer’s Rank (Sto) model.

| **Data Set** | $\tau$ (95% CI) | $\phi$ (95% CI) |
| --- | --- | --- |
| BBC Total (UK) | 0.007 (0.01,0.01) | 0.036 (0.03,0.04) |
| BBC Under 18 (UK) | 0.007 (0.01,0.01) | 0.356 (0.24,0.57) |
| BBC Education (UK) | 0.007 (0.01,0.01) | 0.107 (0.08,0.14) |
| BBC 18-30 (UK) | 0.007 (0.01,0.01) | 0.045 (0.04,0.05) |
| BBC Employed (UK) | 0.007 (0.01,0.01) | 0.037 (0.04,0.04) |
| BBC 30-60 (UK) | 0.007 (0.01,0.01) | 0.037 (0.04,0.04) |
| BBC NEET (UK) | 0.006 (0.01,0.01) | 0.076 (0.06,0.09) |
| BBC 60-100 (UK) | 0.006 (0.01,0.01) | 0.086 (0.07,0.11) |
| Census (E) | 0.007 (0.01,0.01) | 0.2 (0.2,0.2) |
| BBC Total (E) | 0.006 (0.01,0.01) | 0.046 (0.04,0.05) |
| Census (S) | 0.004 (0,0) | 0.281 (0.26,0.3) |
| BBC Total (S) | 0.003 (0,0) | 0.193 (0.16,0.24) |
| Census (W) | 0.003 (0,0) | 0.298 (0.27,0.33) |
| BBC Total (W) | 0.003 (0,0) | 0.279 (0.21,0.37) |
| Census (NI) | 0.002 (0,0) | 0.851 (0.67,1.06) |
| BBC Total (NI) | 0.003 (0,0) | 0.845 (0.48,1.69) |

**Table 14:** Posterior estimates for Impedance (Imp) model.

| **Data Set** | $\phi$ (95% CI) |
| --- | --- |
| BBC Total (UK) | 0.22 (0.21,0.23) |
| BBC Under 18 (UK) | 0.268 (0.21,0.36) |
| BBC Education (UK) | 0.306 (0.25,0.38) |
| BBC 18-30 (UK) | 0.332 (0.3,0.37) |
| BBC Employed (UK) | 0.251 (0.24,0.27) |
| BBC 30-60 (UK) | 0.262 (0.25,0.28) |
| BBC NEET (UK) | 0.319 (0.27,0.39) |
| BBC 60-100 (UK) | 0.361 (0.3,0.44) |
| Census (E) | 0.268 (0.27,0.27) |
| BBC Total (E) | 0.228 (0.22,0.24) |
| Census (S) | 0.369 (0.34,0.4) |
| BBC Total (S) | 0.514 (0.39,0.68) |
| Census (W) | 0.394 (0.35,0.44) |
| BBC Total (W) | 0.952 (0.64,1.44) |
| Census (NI) | 1.133 (0.89,1.42) |
| BBC Total (NI) | 2.388 (1.08,6.06) |

